# Evaluating Visual Photoplethysmography Method

**DOI:** 10.1101/2021.10.18.21265118

**Authors:** Luis Felipe de Deus, Nikhil Sehgal, Debjyoti Talukdar

**Affiliations:** Department of AI Research, Vastmindz Limited, London, United Kingdom; Department of Medical Research, Armenian Russian International University “Mkhitar Gosh”

**Keywords:** Remote Photoplethysmography, Stress Recognition, Heart Rate Variability

## Abstract

Regular monitoring of common physiological signs, including heart rate, blood pressure, and oxygen saturation, can be an effective way to either prevent or detect many kinds of chronic conditions. In particular, cardiovascular diseases (CVDs) are a worldwide concern. According to the World Health Organization, 32% of all deaths worldwide are from CVDs. In addition, stress-related issues cost $190 billion in healthcare costs per year. Currently, contact devices are required to extract most of an individual’s physiological information, which can be uncomfortable for users and can cause discomfort. However, in recent years, remote photoplethysmography (rPPG) technology is gaining growing interest, which enables contactless monitoring of the blood volume pulse signal using a regular camera, and ultimately can provide the same physiological information as a contact device. In this paper, we propose a benchmark comparison using a new multimodal database consisting of 56 subjects where each subject was submitted to three different tasks. Each subject wore a wearable device capable of extracting photoplethysmography signals and was filmed to allow simultaneous rPPG signal extraction. Several experiments were conducted, including a comparison between information from contact and remote signals and stress state recognition. Results have shown that in this dataset, rPPG signals were capable of dealing with motion artifacts better than contact PPG sensors and overall had better quality if compared to the signals from the contact sensor. Moreover, the statistical analysis of the variance method had shown that at least two HRV features, NNi 20 and SAMPEN were capable of differentiating between Stress and Non-Stress states. In addition, three features, IBI, NNi 20, and SAMPEN were capable of differentiating between tasks relating to different levels of difficulty. Furthermore, using machine learning to classify a ‘stressed’ or ‘unstressed’ state, the models were able to achieve an accuracy score of 83.11%.

## INTRODUCTION

There is growing interest in technologies related to remote patient monitoring (RPM) solutions, an interest that has largely piqued as of late amid the COVID-19 pandemic. Furthermore, these technologies can be used to monitor several disorders. Cardiovascular disease (CVD) is a group of disorders related to the heart and blood vessels. According to the World Health Organization [1], 32% of all deaths worldwide are attributed to CVDs. The most common health issues include the risk of a heart attack, stroke, and heart failure.

In addition, stress-related issues place a significant burden on the global healthcare system. Data from the American Institute of Stress [2] shows that 83% of US workers suffer from work-related stress. As a result of work stress, US businesses lose up to $300 billion yearly, and work-related stress issues cause up to 120,000 deaths and result in $190 billion in healthcare costs per year.

RPM solutions are being developed for a multitude of customers of varying ages and varying health conditions of patients are efficiently being monitored outside traditional settings with increased accessibility to regular physiological assessments. Regular physical evaluations of a patient’s health allow the capture of longitudinal data that can be used to provide a highly personalized level of care and has the potential to predict the onset of severe health problems.

Well-established methods for capturing physiological data include the use of electrocardiograms (ECG) or photoplethysmography (PPG), both of which require the use of contact sensors. These methods, through their physiological signals, have the ability and the potential to measure several different physiological parameters such as: heart rate (HR), respiration rate, oxygenation, and mental stress.

One of the main parameters used in these studies is Heart-rate Variability (HRV), which can be accessed through the time difference between two peaks of the signal, either the ECG with the R part of PPG with the systolic peak, in both cases can simply be named the R-R interval.

Since HRV can be deduced from both methods and it is well-established that HRV is an accurate non-invasive proxy to measuring changes in the autonomic nervous system (ANS) and thus accesses the patient’s psychological information. Kim et al. [3], investigated the assumption of correlating HRV and stress levels. Their findings show that HRV features changed in response to stress-induced by different methods. HRV characteristics change in association with low parasympathetic activity, which is characterized by a decrease in the high-frequency band (0.16 Hz - 0.4 Hz) and an increase in the low-frequency band (0.04 Hz - 0.15 Hz).

HRV can also be used to measure potential cardiovascular issues. Although contact-based methods are noninvasive and can capture valuable information about an individual’s health & wellness, they can irritate those with sensitive skin, and such physical devices to capture ECG or PPG signals may not be easily accessible.

Alternatively, researchers have recently introduced the Remote Photoplethysmography (rPPG) technique, which is a low-cost, non-contact method and an alternative solution for measuring the same parameters as the PPG signal in a contact-less way. Since it is a method that can be performed on any consumer technology device with an embedded camera, its ease of use makes it an attractive addition to the suite of RPM solutions.

The information acquired through rPPG reflects the variations of blood volume in skin tissue which is modified by cardiac activity. The reflection of light is influenced by the change in the volume of blood and of the movement of the wall of blood vessels; this phenomenon is visible within frame-to-frame changes of a red, green, and blue (RGB) camera. There are, however several challenges when attempting to retrieve an optimal rPPG signal. Distortion to clean rPPG signals mostly arises from low illumination, significant head movement, a camera’s frame rate, and its resolution.

rPPG methods are usually carried out using a four -step-methodology, which can be summarized as Frame- to-frame extraction, Region of Interest (ROI) detection, signal processing, and vitals estimation. First, the video files are usually separated into several frames, and the amount of frames in a certain period is denoted as frame rate, measured in frames per second (FPS). ROI detection is performed by detecting face regions in each video frame, this process is commonly used with face tracking and landmark detection algorithms such as the Viola-Jones method [4]. Once the ROIs are selected, pixel intensity components are extracted; those components are in the RGB color space. In addition, the RGB components are spatially averaged over all pixels in the ROI to yield a red, blue, and green component for each frame and form the raw signals.

Next, a signal processing stage is applied, also known as the ‘rPPG Core’. This has been the object of various studies in the last decade, resulting in multiple methods which seek to extract reliable rPPG signals from RGB components. Some rely on Blind Source Separation (BSS) methods, which can retrieve information by de-mixing raw signals into different sources.

Principal Component Analysis (PCA)-based and Independent Component Analysis (ICA)-based, which use different criteria to separate temporal RGB traces into uncorrelated or independent signal sources, are some of the used techniques.

ICA separates the pulsatile signal from noise by minimizing the Gaussianity within the de-mixed signal. Mc Duff et al. [5] have used the JADE implementation of ICA to recover source signals from the observations, maximizing the non-Gaussianity within the sources. However, the experiment uses a novel digital single-lens reflex (DSLR) camera, capable of capturing five color channels: red, green, blue, cyan, and orange (RGBCO). On the other hand, Lewandowska et al. [6] obtained the rPPG signal using a 640 × 480 pixels RGB camera, the chosen method was through PCA. The authors then compared the PCA results with FastICA as well as the pulse rate obtained with the ECG ground truth.

Other authors have tried to improve the quality of the signal by changing the color space to a chrominance-based domain. Haan and Jeanne [7], in their work, have proposed a chrominance-based method (CHROM) to extract the rPPG signal by assuming a standardized fixed skin-color tone, where it is assumed to be the same for every-one under white light.

In a recent study, Wang et al. [8] introduced a new alternative to process RGB components into an rPPG signal, called the “plane-orthogonal-to-skin” (POS) algorithm. The main idea of the algorithm is to filter out intensity variations by projecting RGB components on a plane orthogonal to a determined normalized skin tone vector. As a result, a 2-D signal referent to the projections is obtained and then combined into a 1-D signal which is one of the input signal dimensions that is weighted by an alpha parameter. The alpha parameter is the quotient of the standard deviations of each signal.

In this article, we aim to evaluate the efficacy of our rPPG method using the UBFC-Phys public database [9]. The advantage of this dataset over other datasets that are suitable for rPPG evaluation is that UBFC-Phys contains additional data which can be used for stress and emotion analysis. The dataset also contains videos in which subjects are exhibiting significant head movements, which are a good test of how well our rPPG method can handle such conditions.

The remainder of this paper is structured as follows. In section II, an overview of the methodology is presented, which consists of details about the dataset used in this work, the processing techniques needed to cope with noisy signals, as well as a full description of the proposed rPPG Core method. Furthermore, the metrics that were used to evaluate the performance of this work are also shown in this section. Section III presents the results of our experiments that aim to improve the understanding of this data set. In Section IV we discuss our results and the analyses that seek to support the performance achieved. We conclude the article by summarizing our findings and discussing future work ideas in Section V.

## Materials and Methods

The methodology proposed by this work is divided into different stages that go from data acquisition and signal processing to the evaluation method. The flowchart in Fig.1 shows the proposed method.

**Figure 1:**
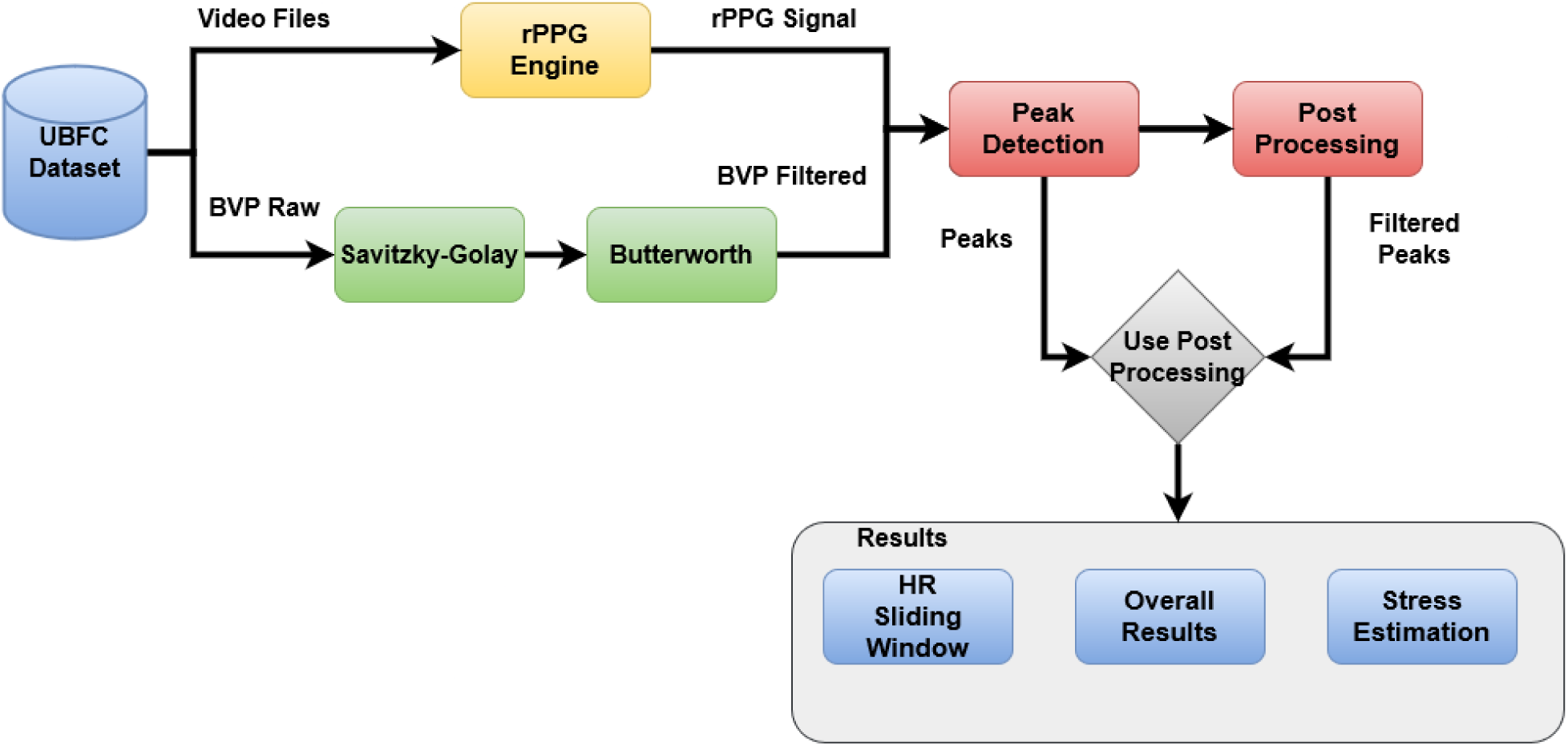
Proposed method - Flowchart.

### Dataset

To evaluate the efficacy of the proposed rPPG methodology, the method was benchmarked against the UBFC-Phys data set [9]. This dataset is a public multimodal dataset and whilst it is dedicated to psychophysiological studies, it contains information that can be used to benchmark general rPPG methods. The dataset contains 56 participants of ages between 19 and 38, with 46 females and 10 males. Each of them followed a 3-step experience that involved a resting task ‘T1’, a speech task ‘T2’, and an arithmetic task ‘T3’. The participants were filmed during each of the three tasks with an EO-23121C camera by Edmund Optics at 35 frames per second and a resolution of 1024 × 1024 pixels. An artificial light source was used to ensure adequate lighting conditions. The participants wore a wristband (Empatica E4) that measured their Blood Volume Pulse signal (BVP), sampled at 64 Hz as well as their ElectroDermal Activity signal (EDA), sampled at 4 Hz. Both the wristband signals and the video file are exactly 3 minutes long.

During the rest of the task, participants were asked to be quiet and not to talk. Therefore, most of the clean signals can be found in the T1 samples. T2 and T3 were interactive tasks and the subjects were randomly assigned to a ‘test’ or ‘ctrl’ group to denote the varying levels of difficulty for each of the speech and arithmetic tasks. 26 subjects were assigned to the “test” group; in this group, they had to endure higher levels of difficulty in comparison to the “ctrl” group, which consisted of 30 individuals, for both tasks. The researcher intended to collect physiological responses that could be well indicative of stress and emotion for the ‘test’ group. Further details of the dataset can be found in the UBFC-Phys original paper [10].

### BVP Processing

Since the BVP signals were obtained using a wrist-band, the body movement performed by the individual can lead to poor signal quality. In an attempt to prove it, a processing stage was performed. Firstly, the BVP signal was re-sampled from its original sampling frequency (64 Hz) to the sampling frequency of our proposed method (30.30 Hz).

Once the BVP signal was resampled, digital filtering techniques were applied to improve the signal quality. Firstly, a 4th order and 19 frames Savitzky-Golay filter [11] was used, to smooth the time series through a moving average. In addition, a 2nd order Butterworth band-pass filter [12], with low-cut and high-cut of 0.7 Hz - 7 Hz respectively, was applied to remove noise artifacts.

### rPPG Processing

In this section, we will describe our method in more detail. The purpose of the method is to find an optimal rPPG signal capable of holding the same physiological information as a PPG signal from a contact sensor. The method is described in Fig.2.

**Figure 2:**
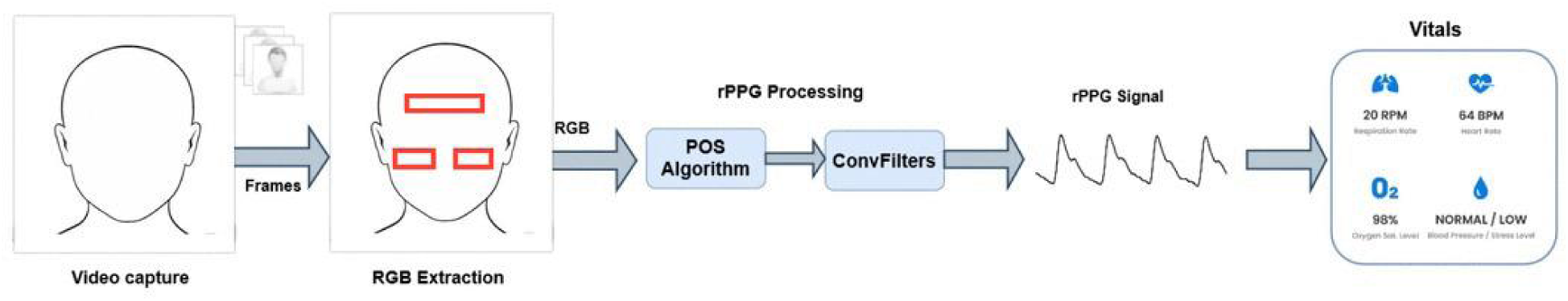
rPPG signal extraction methodology.

The landmark detection algorithm from the OpenCV library [13] was used to extract RGB components from ROIs. This work proposes the use of three ROIs from the forehead, left cheek, and right cheek. Once the raw signal was collected, the rPPG Core, which is composed of the POS algorithm and a filtering stage based on convolutional filters, was applied to extract a clean rPPG signal.

### POS Algorithm

Originally proposed by Wang et al. [8], the POS algorithm seeks to mix RGB channels into a single channel rPPG signal. According to the authors, the input RGB signal channels are mixed on the time interval t as follows:

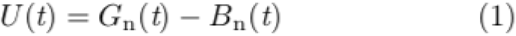

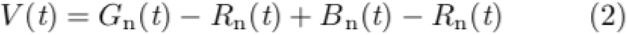

The subscript n represents normalization, representing the instant color values divided by the mean value of the color channel. The rPPG signal on this interval is constructed as denotes Eq.3:

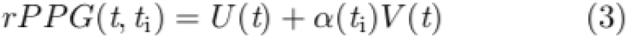

where α is the ratio between the standard deviation of U(t) and V(t) calculated in the interval.

### Convolutional Filter

To clean the signal, this work proposes the use of a convolutional filter (ConvFilter). The ConvFilter will apply the convolution operation of the input single-channel signal ‘s orig’, which is extracted after the POS algorithm, with the template that represents a single heartbeat peak of the same signal. The template is built by averaging segments of the ‘s orig’ signal around the detected peaks in the signal. Since the signal could be a bit noisy, to make peak detection easier, it is recommended to apply a bandpass filter with the pass bandwidth from 0.7 Hz to 7.0 Hz. The cleaner “heart” signal is obtained via convolution Eq.4 or the equivalent correlation Eq.5 with this template t[k]:

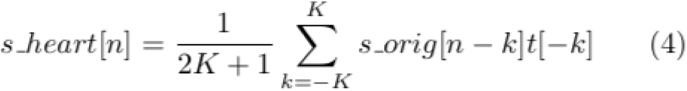

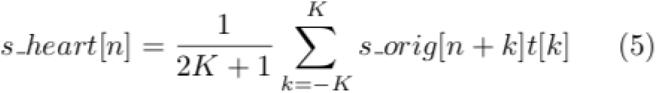

An example of a resulting rPPG signal is shown along with the corresponding BVP GT signal from the dataset in Fig.3.

**Figure 3:**
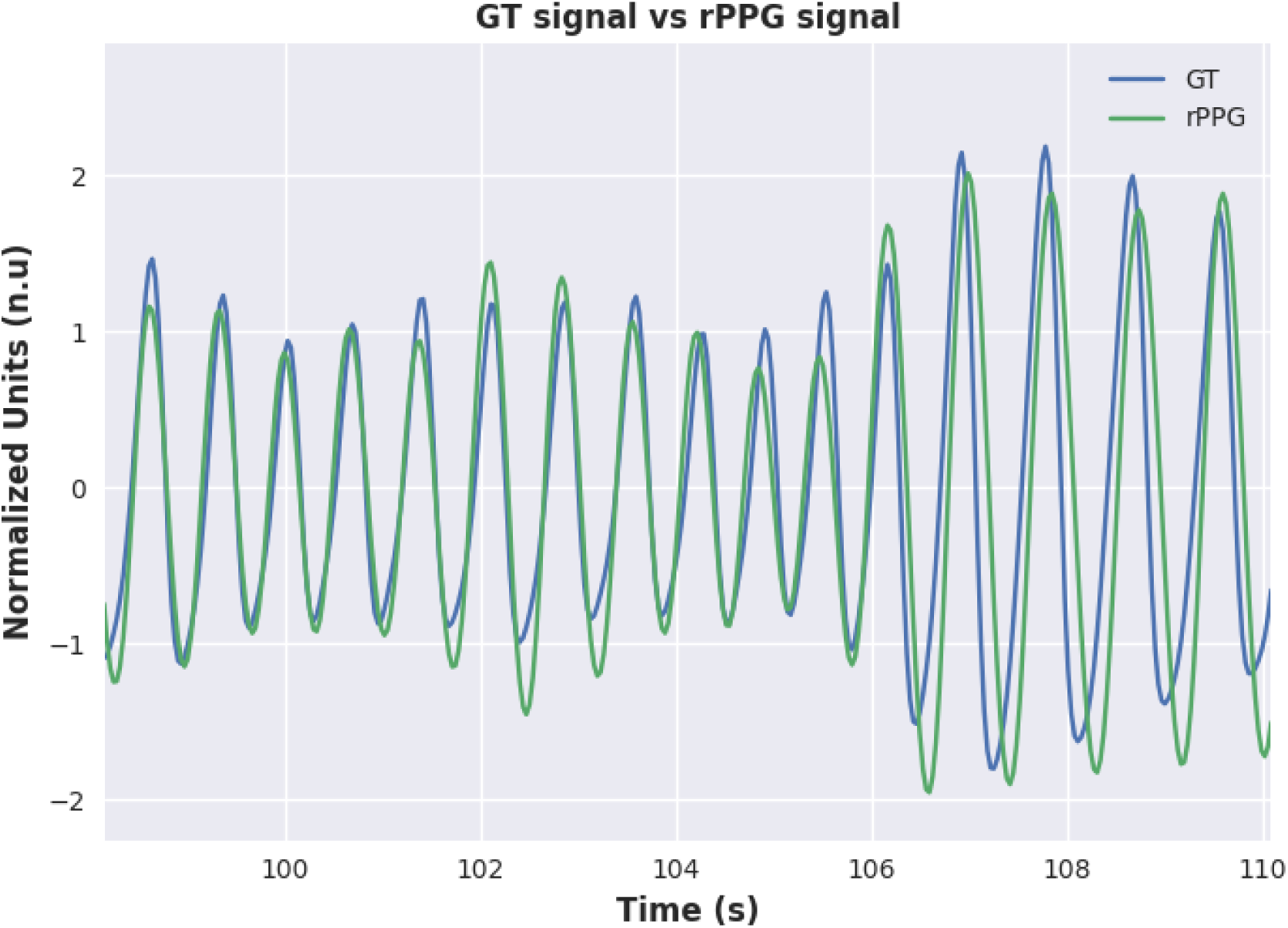
GT Signal vs. rPPG signal.

### Signal-Post Processing

Even though the set of digital filtering techniques, which were applied to the signals, can well remove artifacts and noise issues, it may not be able to improve the signal quality to an acceptable level. Meziati et al. [10] in the original paper, have chosen a strategy to cope with those signals by simply removing them. In our work, we propose a post-processing stage, which seeks to find and remove specific segments of the signal that are damaged in an attempt to avoid removing the entire sample. The procedure is entirely based on detecting corrupted peaks, either peak detected when it should not be or missing peaks. The peak validation process is a set of test rules that are used to find reliable and unreliable peaks either in the BVP or rPPG signals. Firstly, using the original peaks, the IBI signal is calculated. In the first rule, we propose the use of an IBI threshold, in which the IBI signal should be within the range from min IBI to max IBI values that correspond to 210 bpm and 42 bpm of heart rate, respectively. Second, since each peak has its left and right IBI component, for valid peaks, the absolute IBI sequential difference should be less than 0.5 seconds.

The valid peaks are those that have valid both their left and right IBI components. Therefore, the first detected peak in the signal is invalid because it has no left IBI, and the last peak is invalid because it has no right IBI.

Finally, a peak that is just before or after the invalid peak is assumed to be invalid. This processing is applied only once, as a last validation test. In this way, there are two invalid peaks at the beginning and the end of the signal. Moreover, 4 invalid peaks can be expected to appear around 1 invalid IBI in the signal. This rule aims to eliminate isolated peaks surrounded by invalid peaks. Fig.4 presents an example of a post-processing algorithm in use.

**Figure 4:**
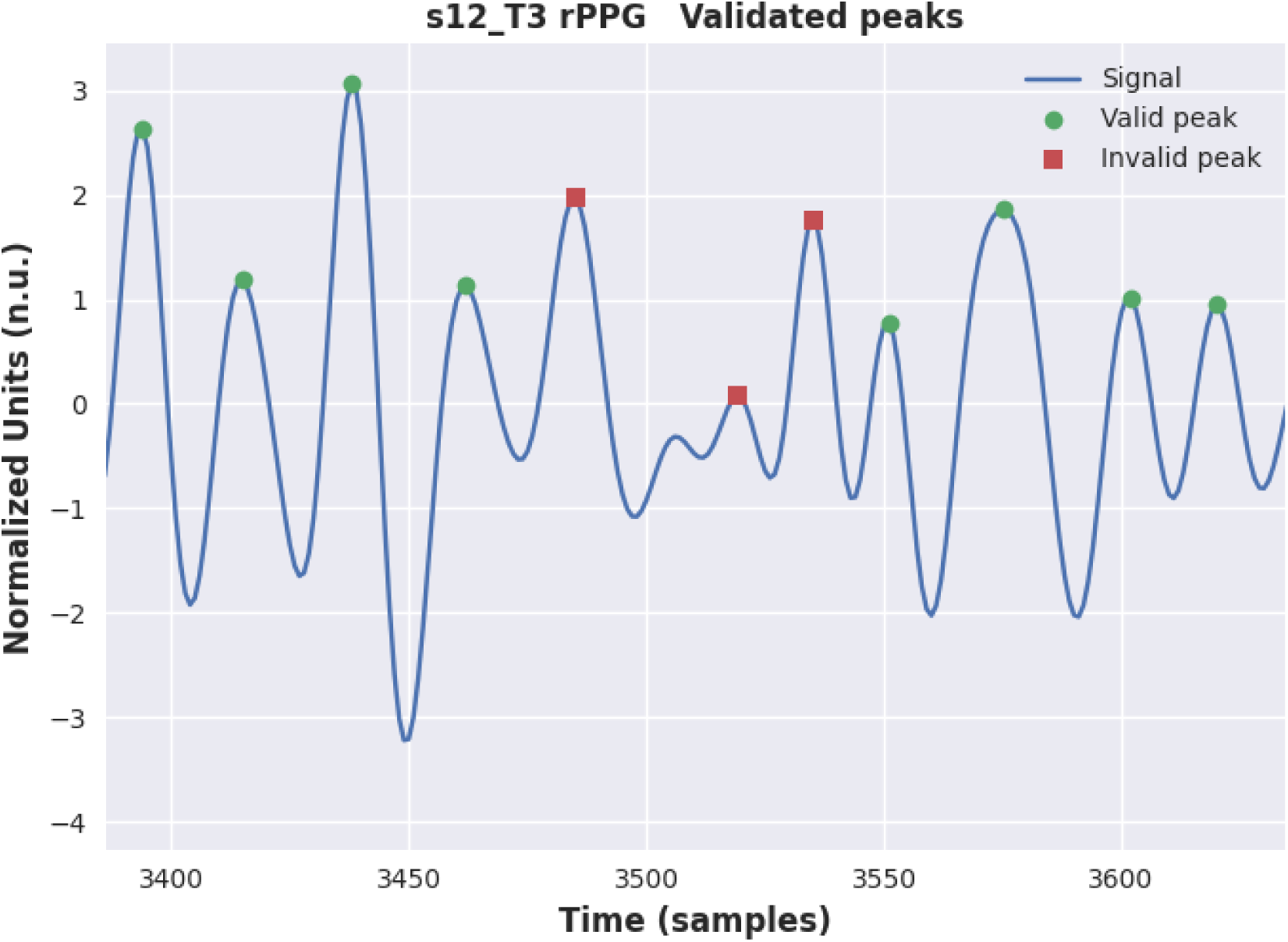
Post-processing algorithm over the rPPG signal.

### Metrics

As stated previously, this work seeks to evaluate our proposed method of remote health screening using rPPG signals extracted from video files. To compare the performance of our method, the BVP signal was used as the “ground truth”. Thus, physiological features were taken from both contact and remote signals and then compared.

The main tool used to calculate those features is the RR Interval, also known as pulse-to-pulse interval, which is the time difference between two peaks in terms of milliseconds (ms) Eq.6. Heart rate (HR), measured in beats per minute (bpm), can be calculated from the RR intervals, Eq.7 shows the definition. In addition, we also compile the following features in the time domain: InterBeat-Interval (IBI) Eq.8, Root Mean Square of Successive Differences between Normal Heartbeats (RMSSD) Eq.9 and the Standard Deviation of Normal to Normal Heartbeats (SDNN) Eq.10. In the frequency domain, we measured the power from the low-frequency band (LF) [0.04; 0.15]Hz and the high-frequency band (HF) [0.15; 0.4]Hz.

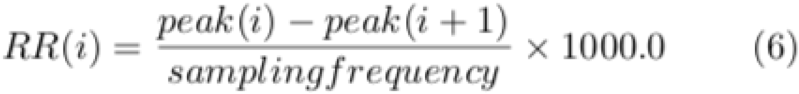

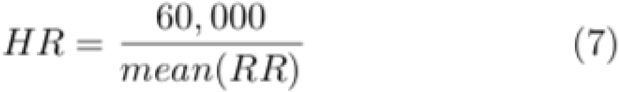

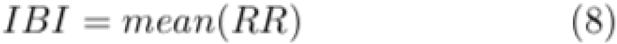

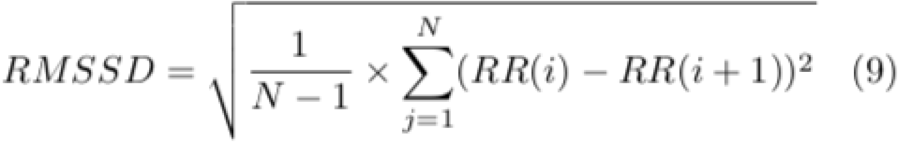

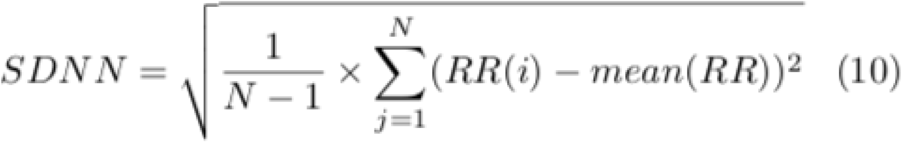

To evaluate the accuracy of our rPPG method against the contact sensor, we measured the mean absolute error (MAE) for each comparative feature described above. For the stress classification task, statistical metrics were used, as described in Eq.11, Eq.12, Eq.13, Eq.14, and Eq.15. We also use the F1-Score Eq.15 to evaluate how well the rPPG method can identify pulsatile peaks in a signal.

The F1-Score is the weighted average of Precision Eq.12 and Recall Eq.13, which the score takes both, false positives and false negatives into account. The concepts of true positives, false positives, and false negatives are entirely based on the position of the pulsatile peaks in the signal.

- **True Positive (TP):** Event in which there was a peak and the algorithm found it correctly.
- **False Positive (FP):** Event in which there was not a peak and the algorithm incorrectly points.
- **False Negative (FN):** Event in which there was a peak and the algorithm has not found it.

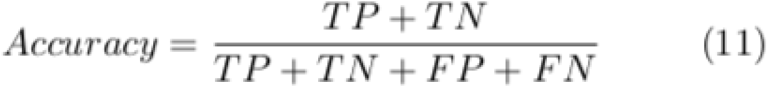

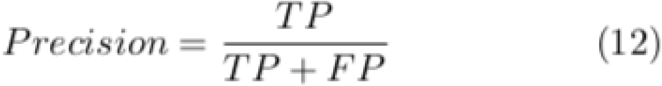

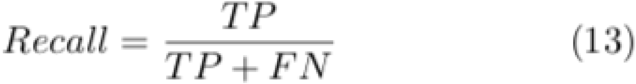

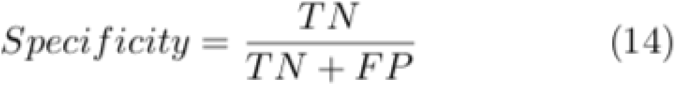

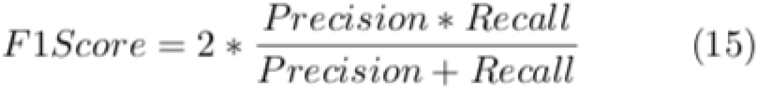

### Stress Experiments

This work also seeks to evaluate how well we can predict whether an individual is under stress or not, just by looking at features of the rPPG signal. In the UBFC-Phys dataset, there are at least three different ways to measure this. Stress classification between stress and unstress, task differentiation between subjects in T1, T2, and T3, and finally group differentiation between test and control.

To conduct the analysis, only HRV features in the time, frequency, and nonlinear domains were used. Moreover, this work does not use the self-reported scores assigned to subjects before and after the experiment. The group separation between stress and non-stress was created in the following way: All T1 samples were labeled as non-stress, T2 and T3 samples were labeled as stress if the sample belongs to the test set otherwise, if the subject belongs to the control set, then this sample is labeled as non-stress.

In our work we used two different approaches for each one of the experiments, using a statistical One-Way Analysis of Variance (ANOVA) test and Machine Learning (ML) models. ML models are often used for classification tasks, in this work we propose the use of HRV features as inputs to those models.

Moreover, ANOVA was used to verify the hypothesis that the groups are separable. ANOVA is a statistical tool that compares means of several samples, this is done by analyzing the variances between the data and within these groups, it is also known as an extension of the t-test for two independent samples to more than two groups [14]. ANOVA test of the hypothesis is based on a comparison of two independent estimates of the population variance [15].

The null hypothesis of the ANOVA test represents that there is no difference between the groups, to reject this hypothesis, the p-value resulting is observed and compared with a certain threshold, usually a significance level of 0.05. In other words, if the resulting p-value is below (p-value < 0.05), it rejects the hypothesis and it is possible to conclude with 95% confidence that there are differences between the means of the group, thus the groups are separable.

However, to use the ANOVA test, a normally distributed data set is required. To test whether a feature follows a normal distribution or not, the Shapiro-Wilk test [16] was applied. The Shapiro-Wilk test is based on the correlation between the data and the corresponding normal scores [17]. The null hypothesis of the Shapiro-Wilk test is that the feature is normally distributed, which is represented by a p-value above 0.05 (p-value > 0.05).

Even though ANOVA is a powerful tool that indicates that at least one group differs from the other groups, the method does not show which particular group differs or if there is more than one. Thus, ANOVA is often equipped with a specific test that compares the two means between the pairs or groups, also known as pairwise comparisons. In this work, Tukey’s test was applied [18].

## Results

In this study, video files from the UBFC data set [9] were used to extract physiological information from individuals. Each one of the videos was processed using our proposed method described in section II C, which returns the rPPG signal.

The information obtained through the rPPG signal was then compared to that obtained from the BVP signal, which was taken as ground truth.

Firstly, after the rPPG signals were obtained, the post-processing algorithm described in section IID was used to better understand the relationship between the quality of the contact and remote signals. Fig.5 shows the percentage of validated peaks, according to the post-processing algorithm, in each sample per task for both Contact GT and Remote rPPG signals. Moreover, Fig.6 shows the calculated F1 score obtained over the synchronized GT and rPPG peaks.

**Figure 5:**
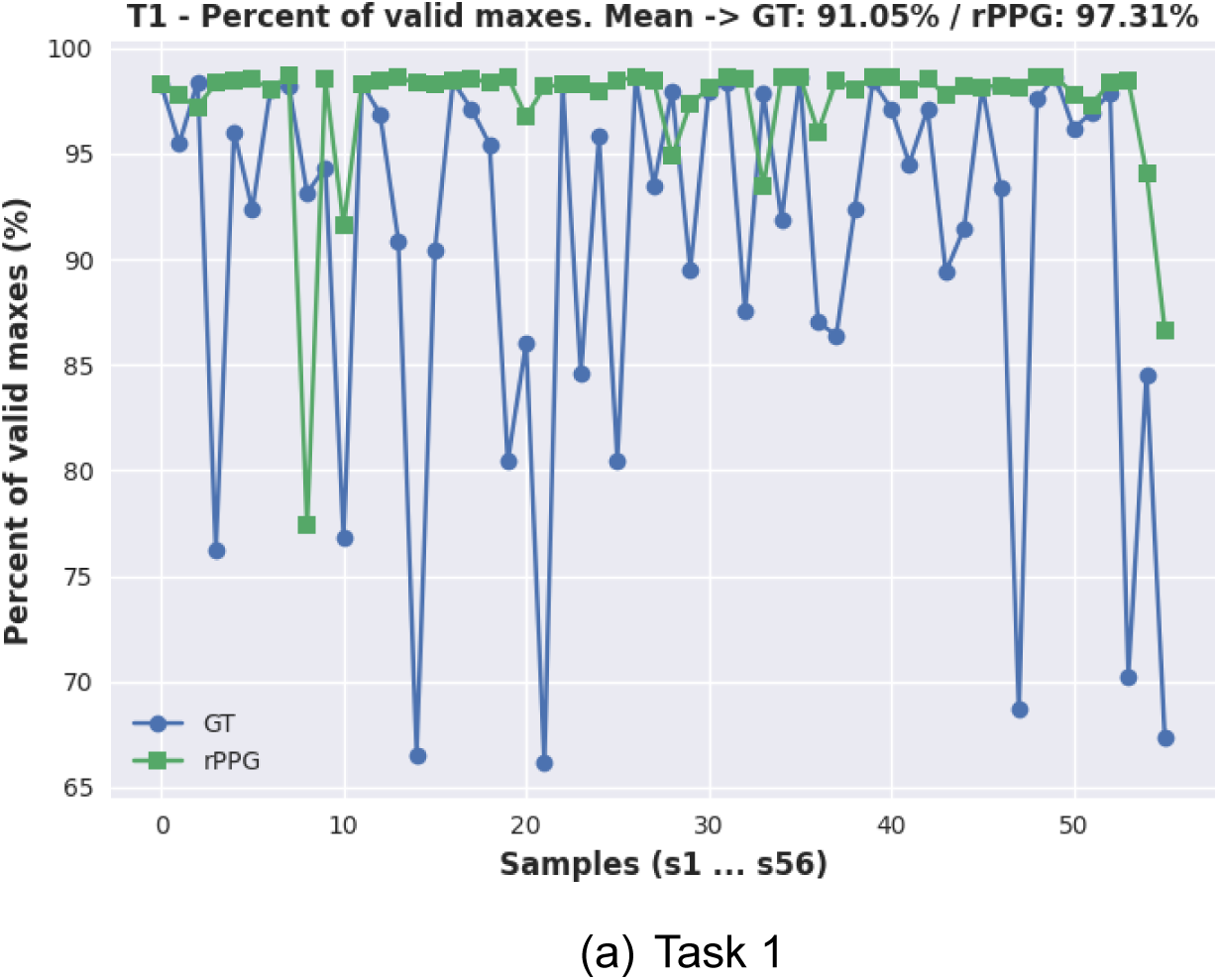
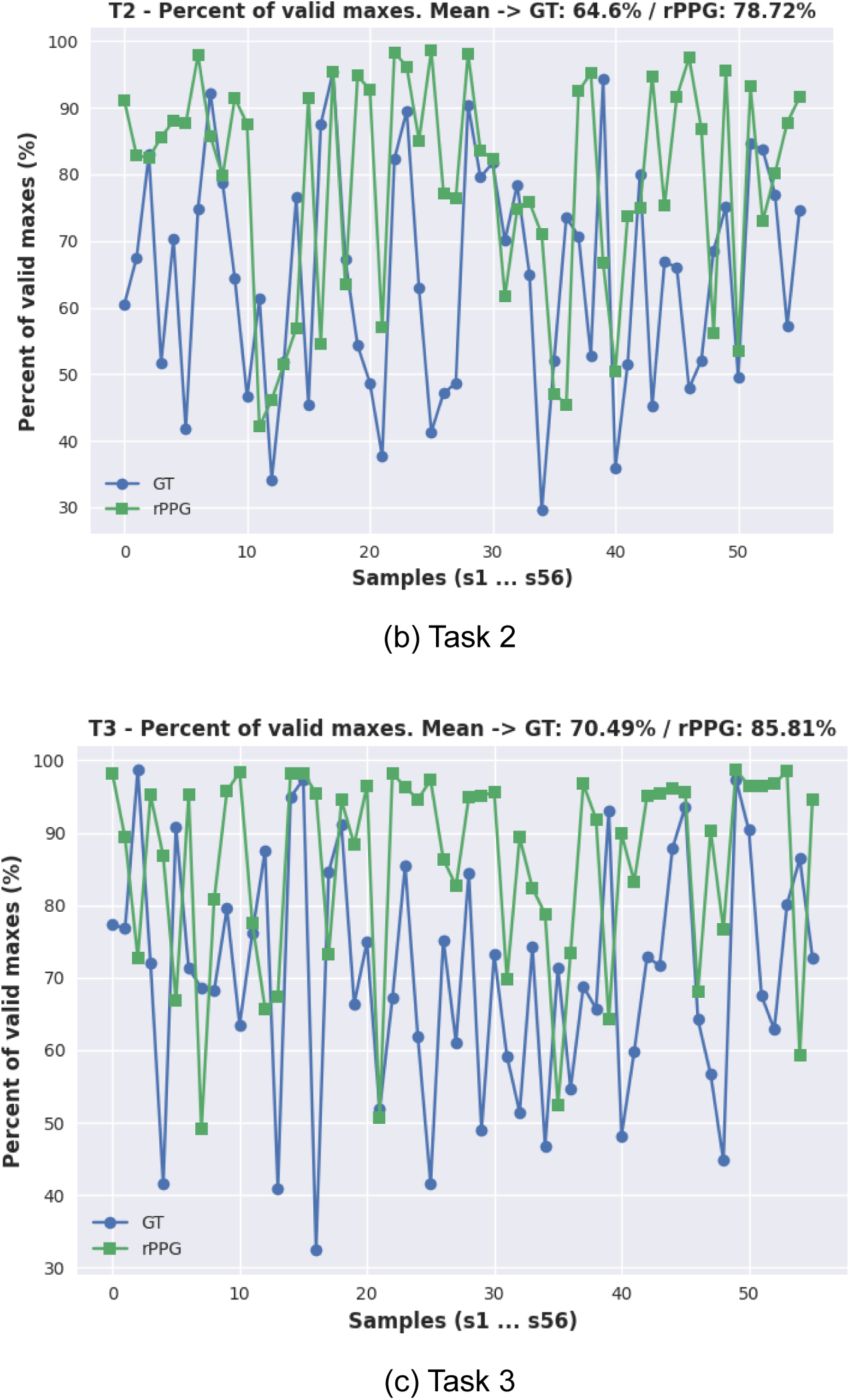
Percentage of valid maxes (peaks) for contact GT / remote rPPG in each sample per task.

**Figure 6:**
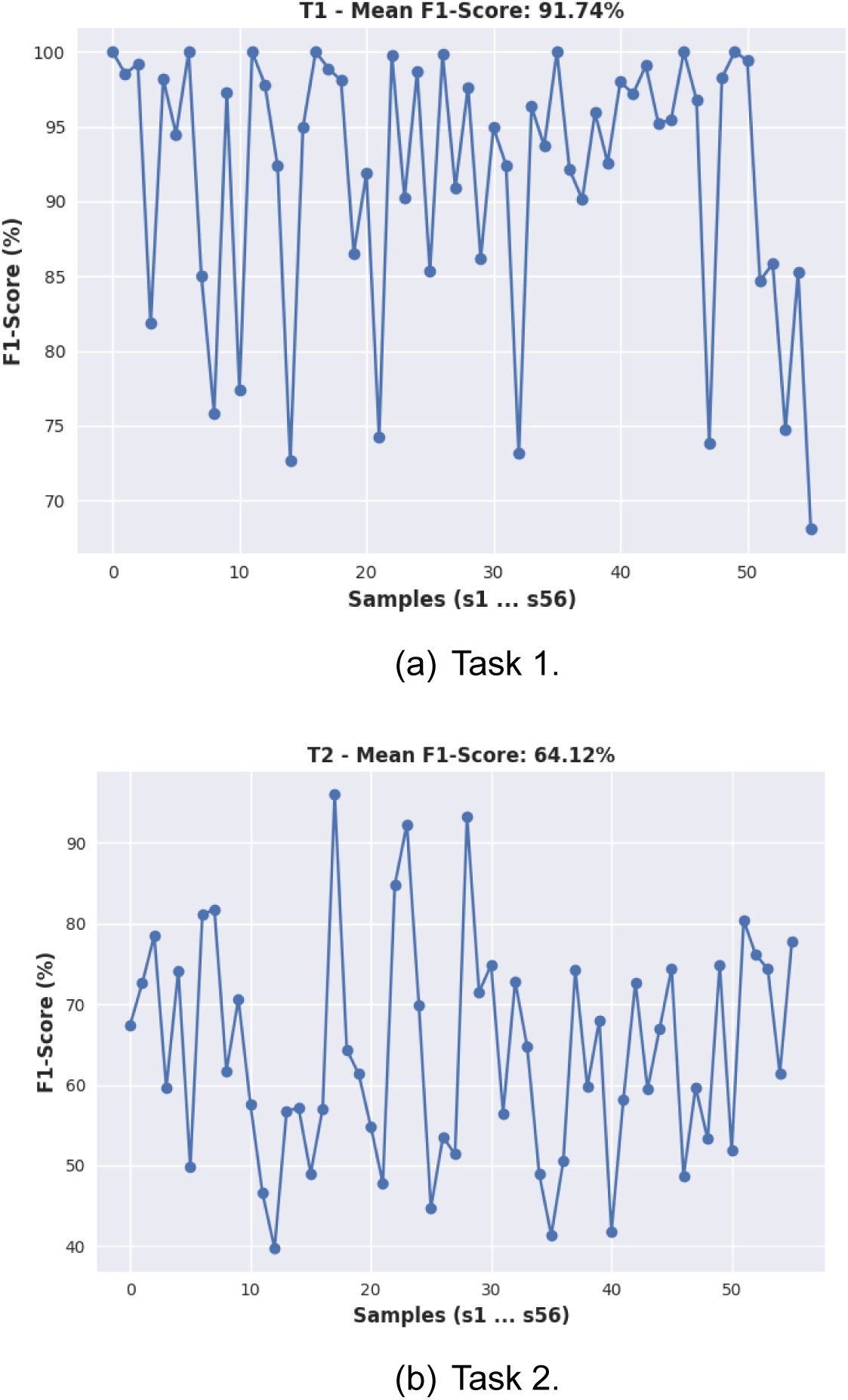
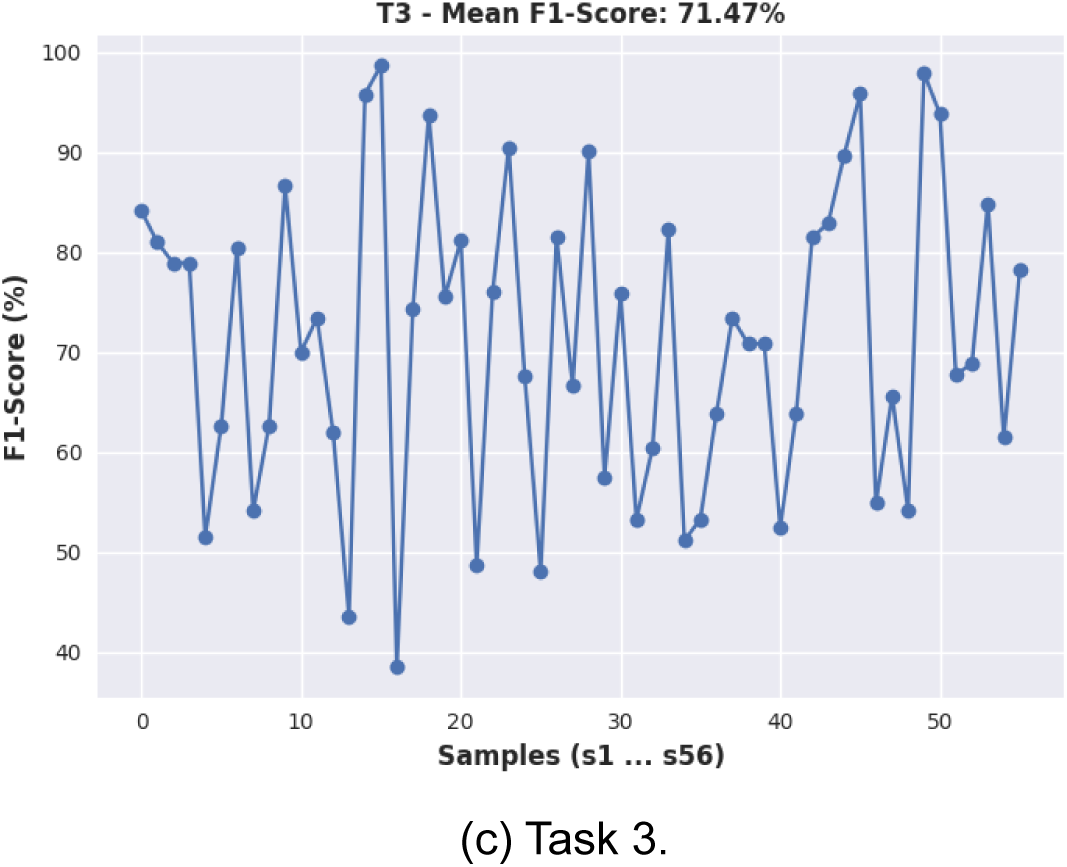
F1-Score percentage obtained from TP, FP, and FN considering Contact and Remote extracted peaks

To extract the heart rate values over time from each subject, a sliding window approach, with a size of 30 seconds and stride of 1 second was used. Fig.7 shows the results comparing HR estimation using the BVP GT and rPPG signals, for three different samples and tasks, in agreement with the original experiment setup. Those results are only using filtering techniques to clean the signal. Moreover, Fig.8 shows the same experiment, as ever, now using the proposed post-processing algorithm.

**Figure 7:**
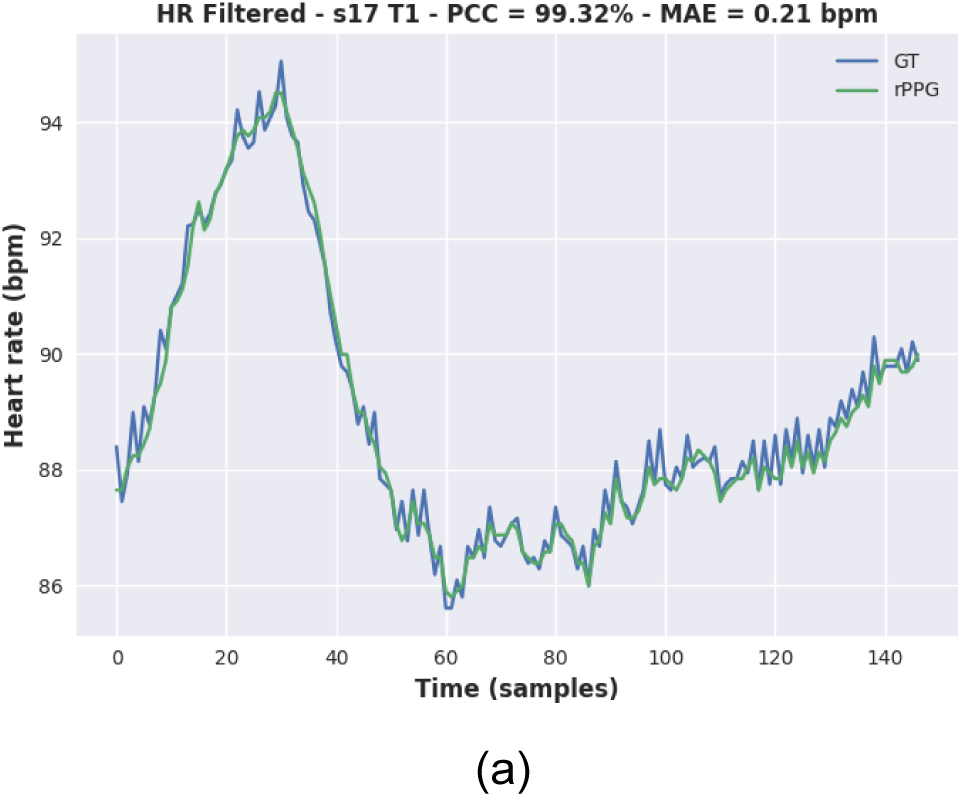
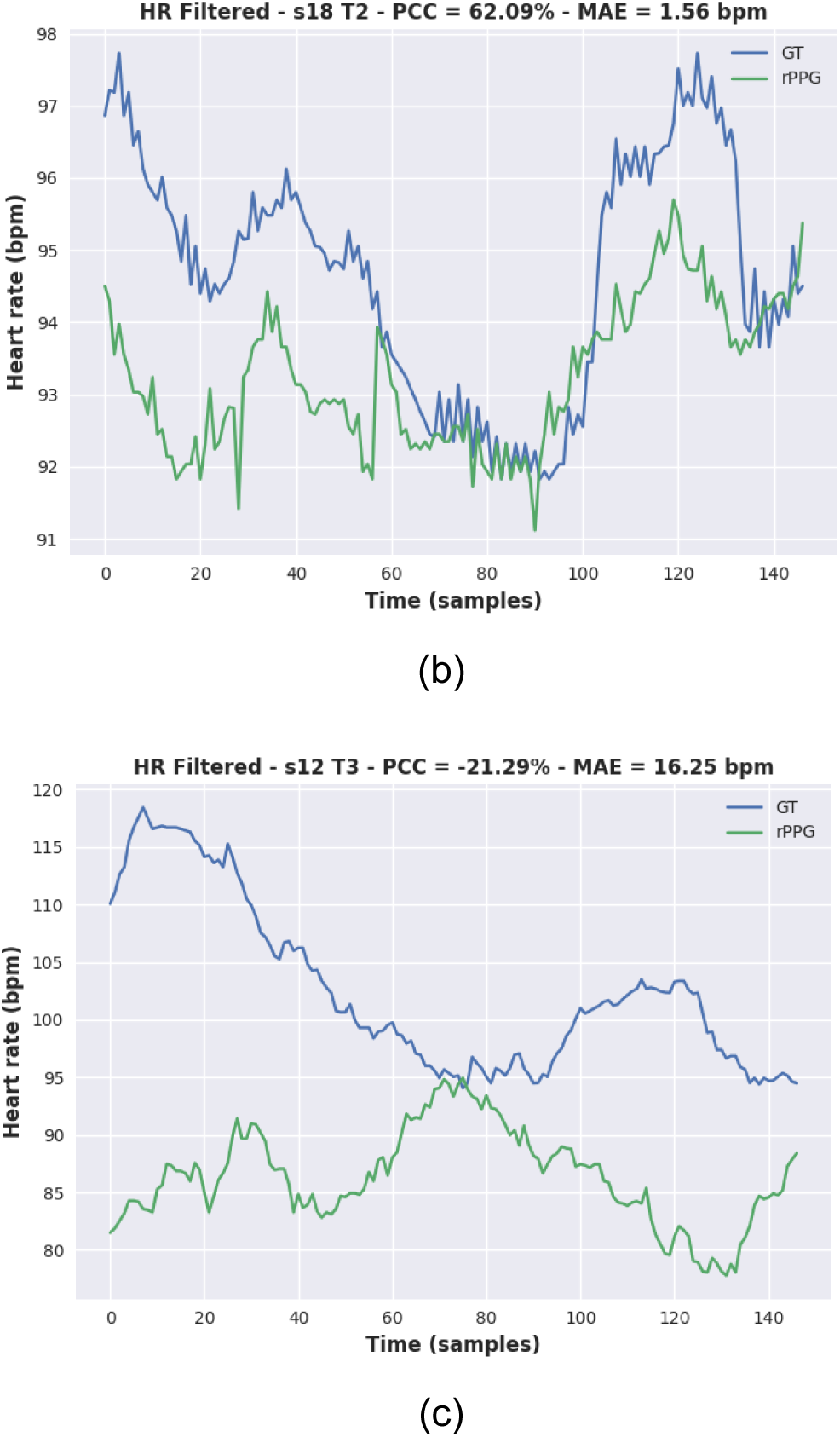
Remote and Contact heart rate signals using 30 seconds sliding window - Results with filtered signals, no post-processing stage.

**Figure 8:**
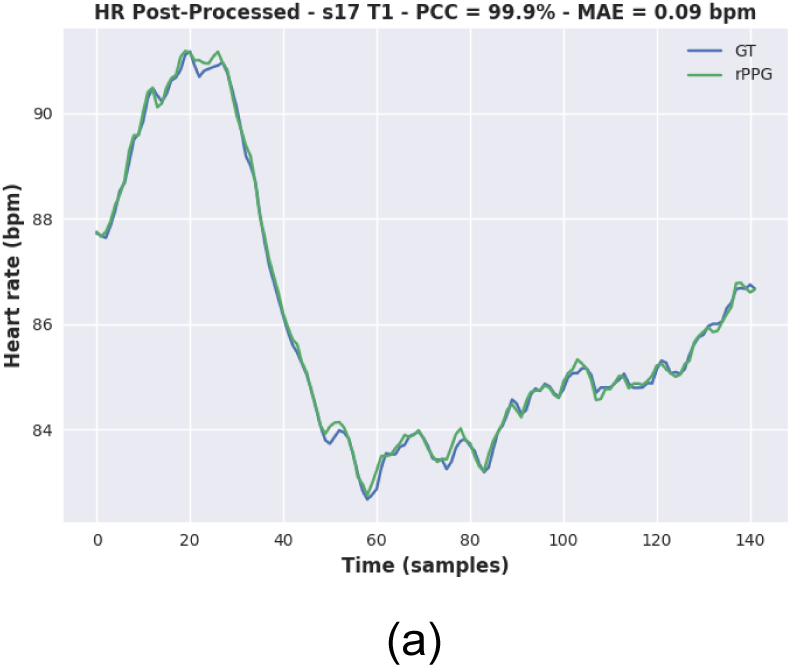
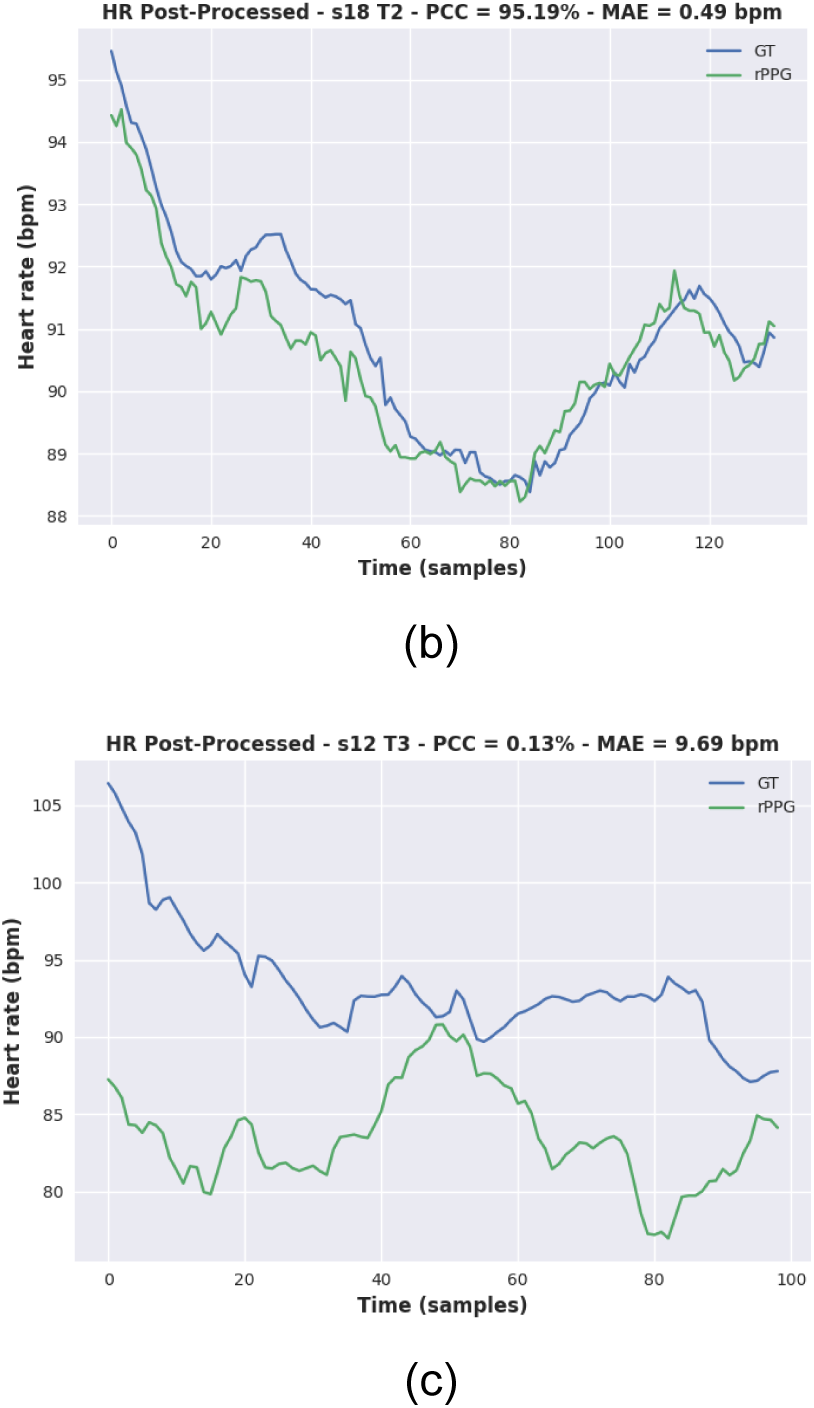
Remote and Contact heart rate signals using 30 seconds sliding window - Results after post-processing algorithm.

To understand the real spatial difference between estimated and ground-truth HR, two plots were created. First, Fig.9 shows the scatter plots for each task, where the x-axis represents the presumed correct value from the contact sensor and the y axis represents the remote estimated heart rate, both calculated using Eq.7. In addition, the best-fit line, as well as the perfect 1:1 line, are also shown to visualize the correlation. The best-fit line was obtained using a first-degree polynomial function using Eq.16, with the following coefficients:

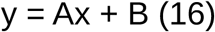

- T1: A: 0.891862, B: 11.74
- T2: A: 0.377204, B: 51.50
- T3: A: 0.661961, B: 27.63

**Figure 9:**
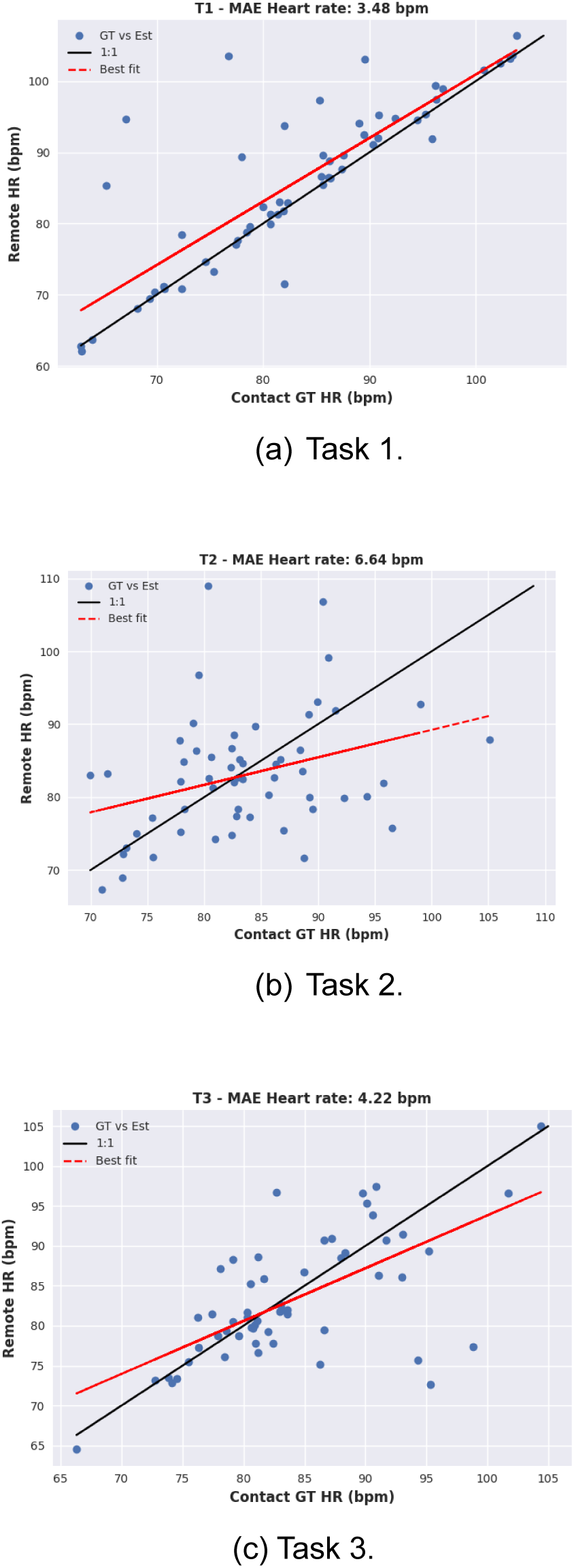
Contact vs. Remote - overall mean heart rate estimation.

Moreover, Fig.10 brings the heart rate error level distribution for each one of the three tasks. The bins correspond, respectively, to

- Error < 5
- 5 ≤ Error < 10
- 10 ≤ Error < 15
- Error ≥ 15

**Figure 10:**
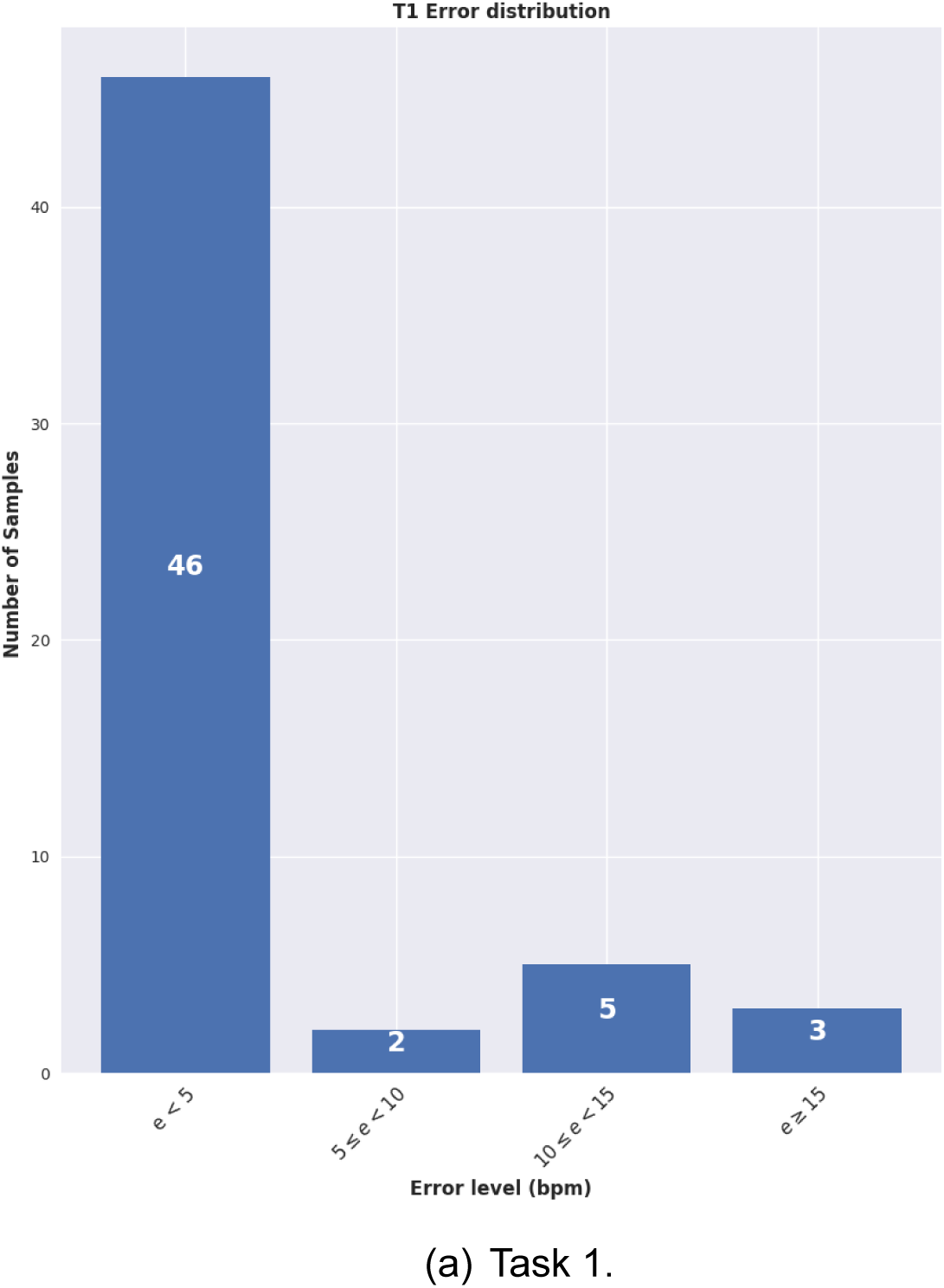
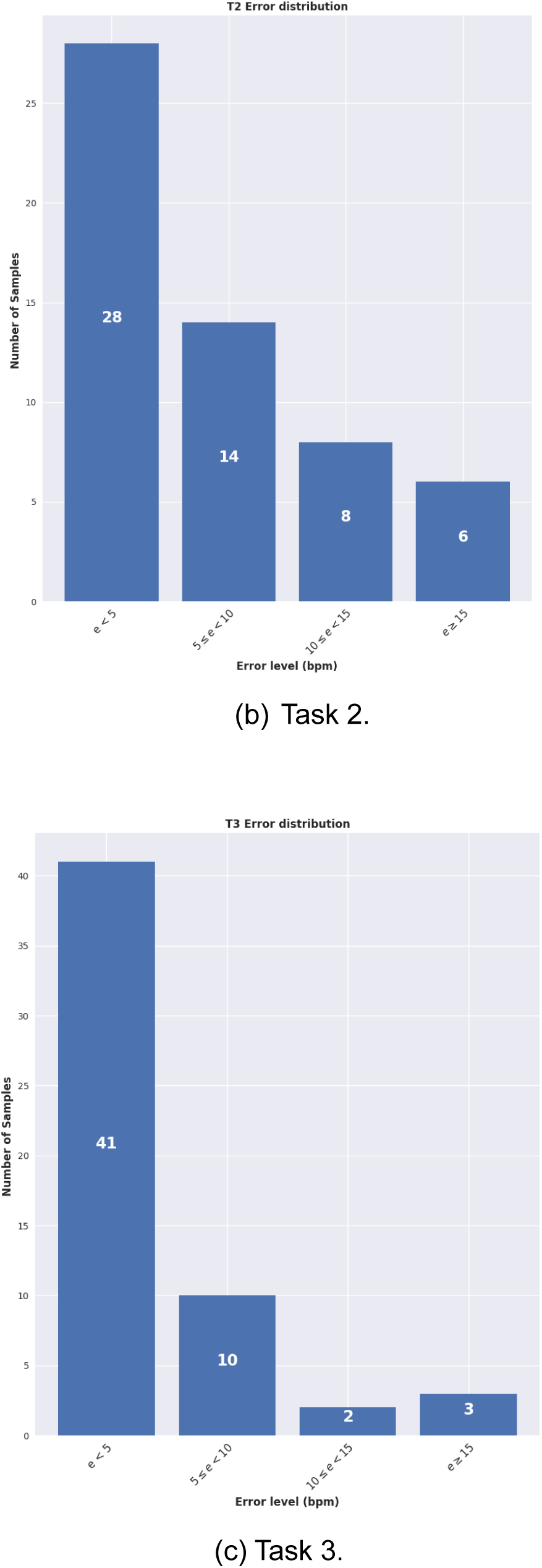
Overall heart rate error level distribution.

It’s clear to see that the UBFC dataset has damaged signals, and these signals can be either one of BVP GT or rPPG signals or both. In contrast with the original article, which proposes exclusion criteria based on a comparative assessment between the rPPG and BVP signals, this work aims to create a more robust approach to identify unreliable signals. This condition must be under the assumption that four scenarios are possible:

a. Acceptable GT and acceptable rPPG.
b. Acceptable GT and unacceptable rPPG.
c. Unacceptable GT and acceptable rPPG.
d. Unacceptable GT and unacceptable rPPG.

These scenarios are apparent in any of the three tasks. Fig.11 shows one example of a signal for each possible scenario.

**Figure 11:**
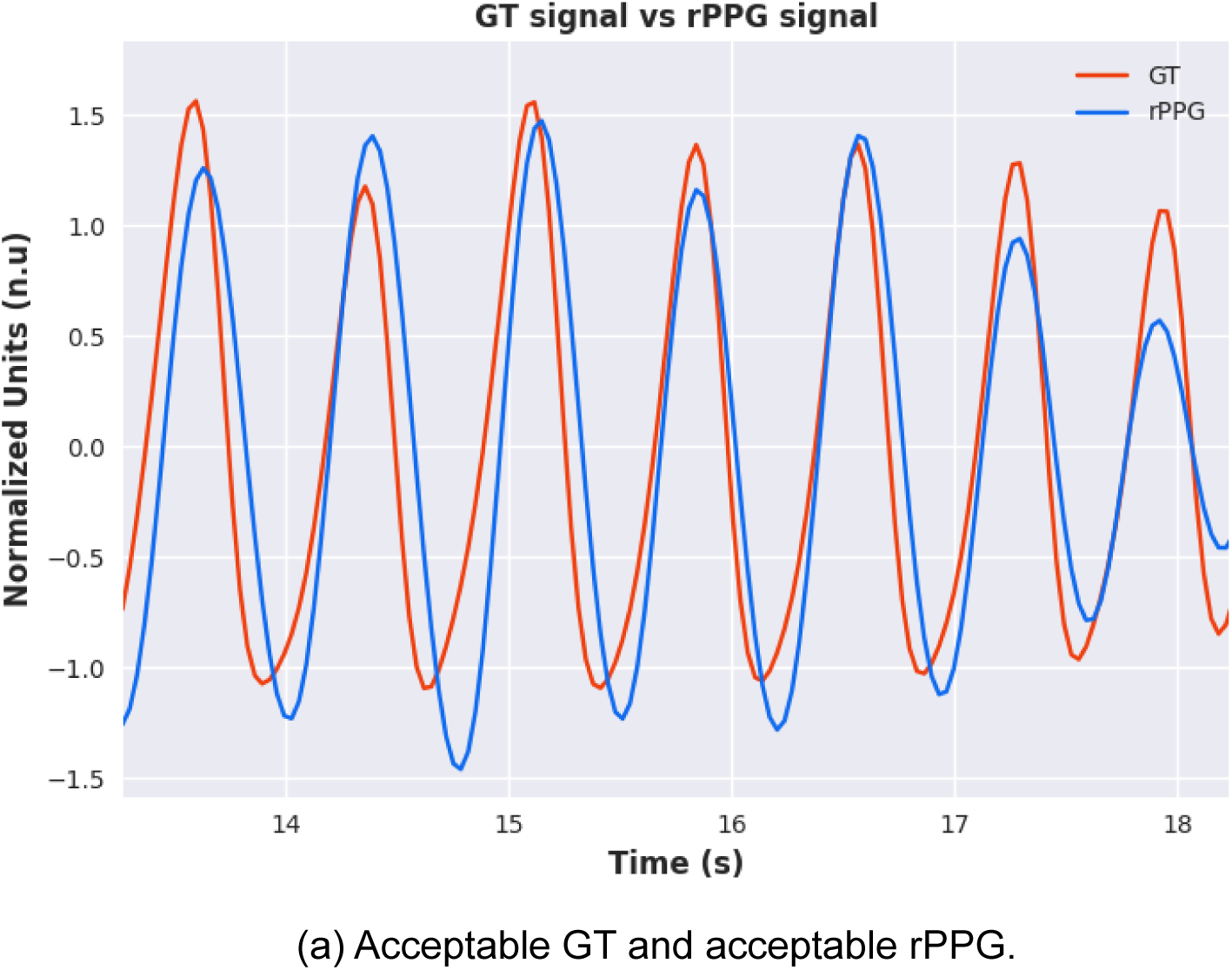
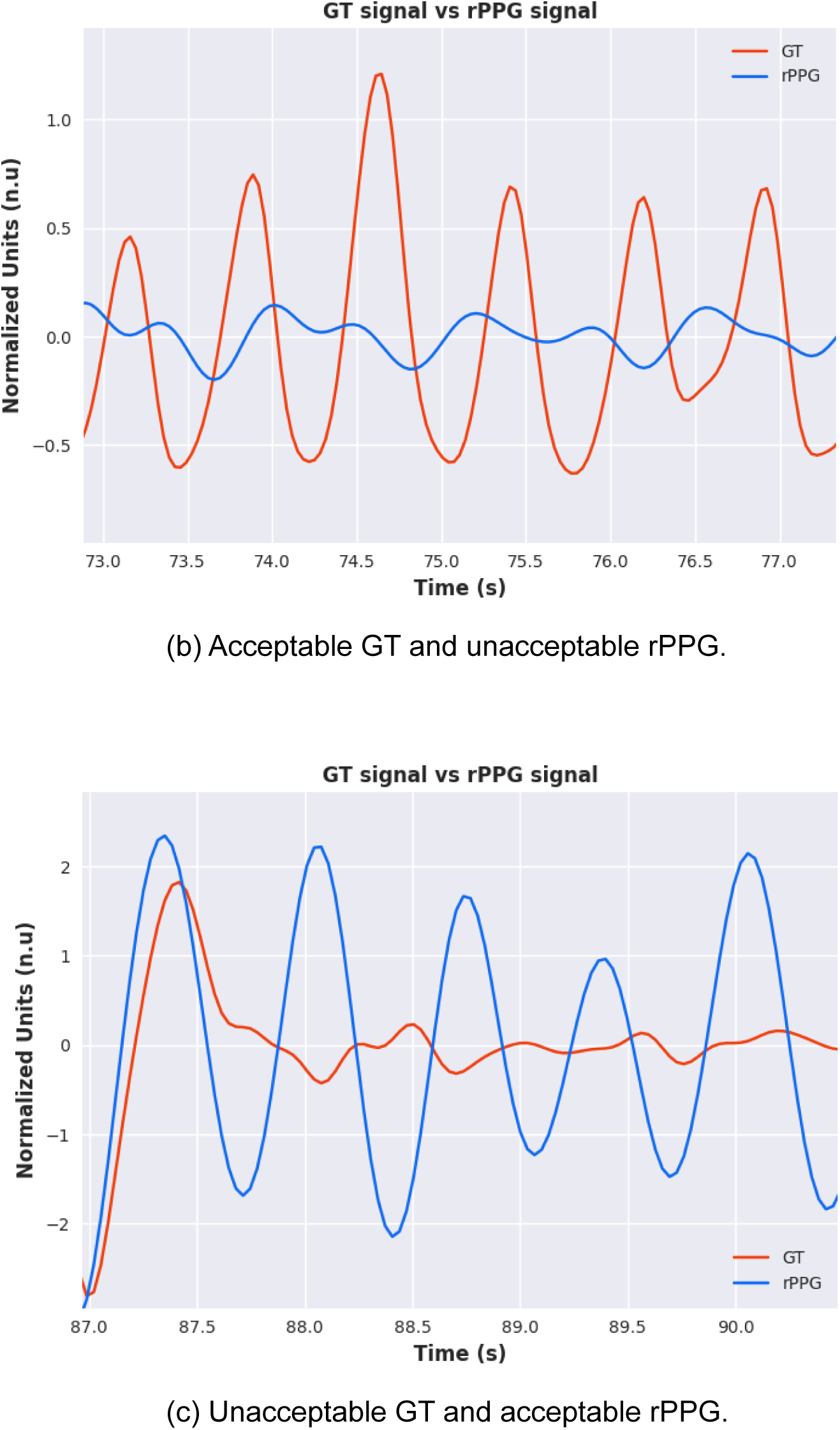
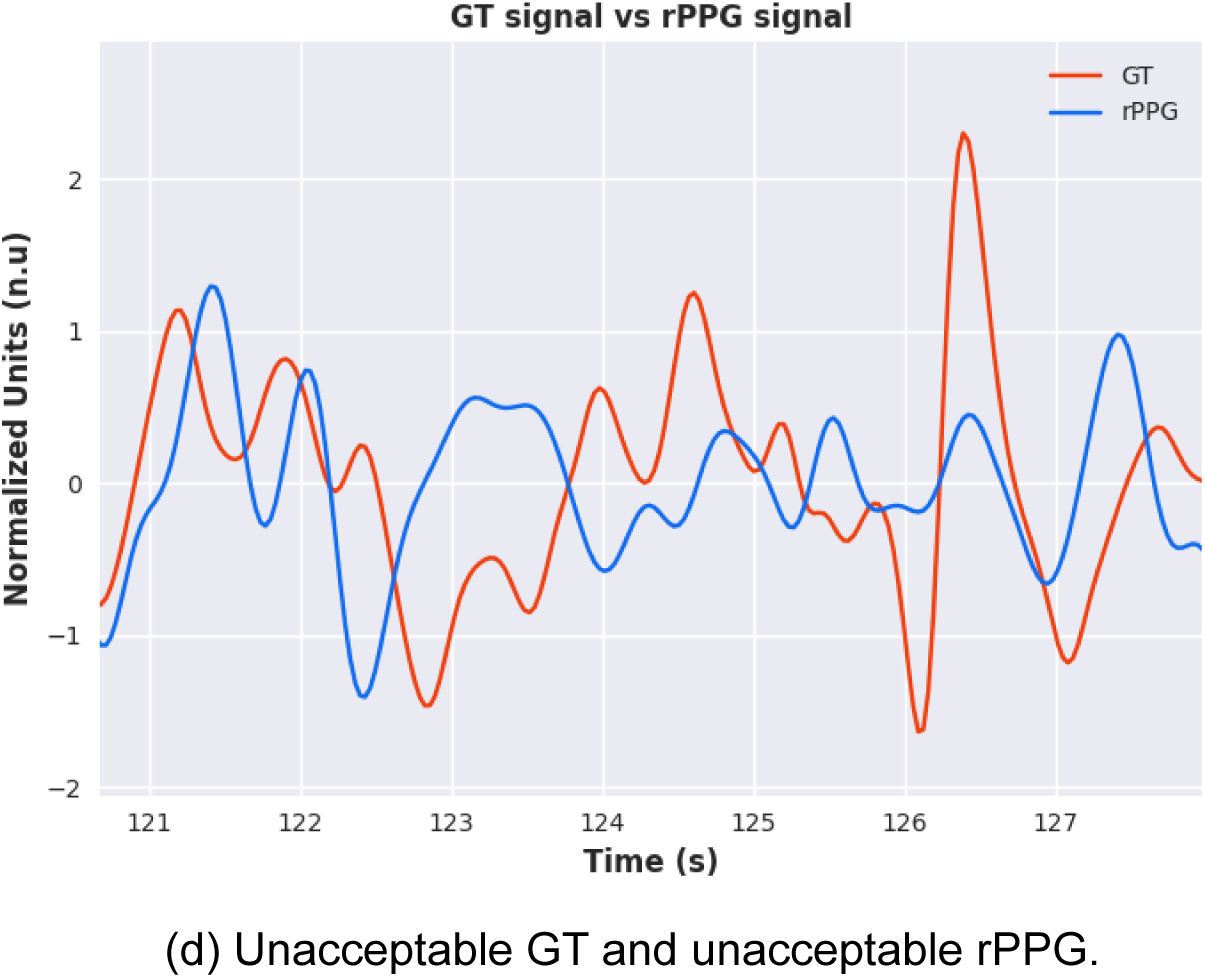
Contact vs. Remote - Signal Quality.

This work looked at several parameters to formulate a fair and robust exclusion criterion solely based on removing damaged GT signals. To reach this goal, the following parameters were investigated.

- Percentage of GT valid peaks;
- F1-Score of valid peaks (uses GT and rPPG peaks);
- Signal-to-noise ratio (SNR);
- Standard deviation of GT heart rate.

Experiments have shown that the most efficient and reliable parameter was the percentage of GT valid peaks. However, the threshold used as the minimum required value to validate the sample has its limitations. Table I shows the results for different thresholds of validated GT peaks.

**Table I:** Overall abs. heart rate error using different exclusion criteria. Validated peaks obtained from GT.

**Evaluation of exclusion criteria - Abs. Error HR (bpm)**
|  | GT Validated Peaks Score |  |  |  |
| --- | --- | --- | --- | --- |
| T1 | TH > 60% | TH > 70% | TH > 80% | TH > 90% |
| no. files | 56 | 52 | 49 | 39 |
| mean value | 3.38 | 3.23 | 3.42 | 2.94 |
| Min value | 0.01 | 0.01 | 0.01 | 0.01 |
| max value | 33.05 | 33.05 | 33.05 | 33.05 |
| **T2** | **TH > 60%** | **TH > 70%** | **TH > 80%** | **TH > 90%** |
| no. files | 34 | 24 | 11 | 4 |
| mean value | 5.58 | 4.99 | 3.72 | 3.81 |
| Min value | 0.00 | 0.00 | 0.37 | 1.28 |
| max value | 26.34 | 26.34 | 8.87 | 5.82 |
| **T3** | **TH > 60%** | **TH > 70%** | **TH > 80%** | **TH > 90%** |
| no. files | 42 | 30 | 16 | 9 |
| mean value | 4.02 | 4.15 | 3.12 | 4.18 |
| Min value | 0.06 | 0.06 | 0.06 | 0.39 |
| max value | 22.31 | 13.69 | 13.69 | 13.69 |

Although using an exclusion criterion based on GT valid peaks can increase the reliability of the results, it is possible to see that the maximum value of all three tasks was considerable. Those large errors can be accepted if the problem was due to suboptimal rPPG signals, which can be understood as those samples in which the rPPG method failed, however, this is not always the case.

Using the samples that survived from the exclusion criterion of GT valid peaks > 60%, each sample that could be a potential outlier was investigated by evaluating the Signal to Noise Ratio (SNR) of the GT signal. Using a threshold of 4 dB, we removed GT signals that fell below this threshold, indicating that this signal was not of sufficient quality for comparative evaluation. Table II shows the results using these two exclusion criteria. Table II: Results after exclusion criteria of GT valid peaks > 60% and SNR > 4dB.

**Table II:** Results after exclusion criteria of GT valid peaks > 60% and SNR > 4dB.

**MAE HR (bpm)**
| **T1** | **This work** |
| --- | --- |
| no. files | 55 |
| mean value | 3.33 |
| min value | 0.01 |
| max value | 33.05 |
| **T2** | **This work** |
| no. files | 22 |
| mean value | 3.08 |
| min value | 0.00 |
| max value | 16.26 |

| T3 | This work |
| --- | --- |
| no. files | 34 |
| mean value | 2.77 |
| min value | 0.06 |
| max value | 22.31 |

Despite applying these techniques to avoid including damaged GT signals in our analysis, it is still evident that damaged GT signals were still left. Therefore, a manual check on the GT signal was performed to avoid damaged signals that neither of the proposed materials was able to filter. We only found additional damaged signals in Task 1 during our manual check. Fig. 12, shows an example of a damaged GT signal with good quality rPPG, and Table III presents the results after applying our exclusion criteria which is based on: Valid GT valid peaks > 60%, SNR > 4 dB, and a manual GT signal check.

**Table III:** Results of this work after proposed exclusion criteria, GT valid peaks > 60%, SNR > 4 dB, and manual GT signal check.

**MAE HR (bpm)**
| T1 | This work |
| --- | --- |
| no. files | 47 |
| mean value | 1.10 |
| min value | 0.01 |
| max value | 4.70 |
| T2 | This work |
| no. files | 22 |
| mean value | 3.08 |
| min value | 0.00 |
| max value | 16.26 |
| **T3** | **This work** |
| no. files | 34 |
| mean value | 2.77 |
| min value | 0.06 |
| max value | 22.31 |

**Figure 12:**
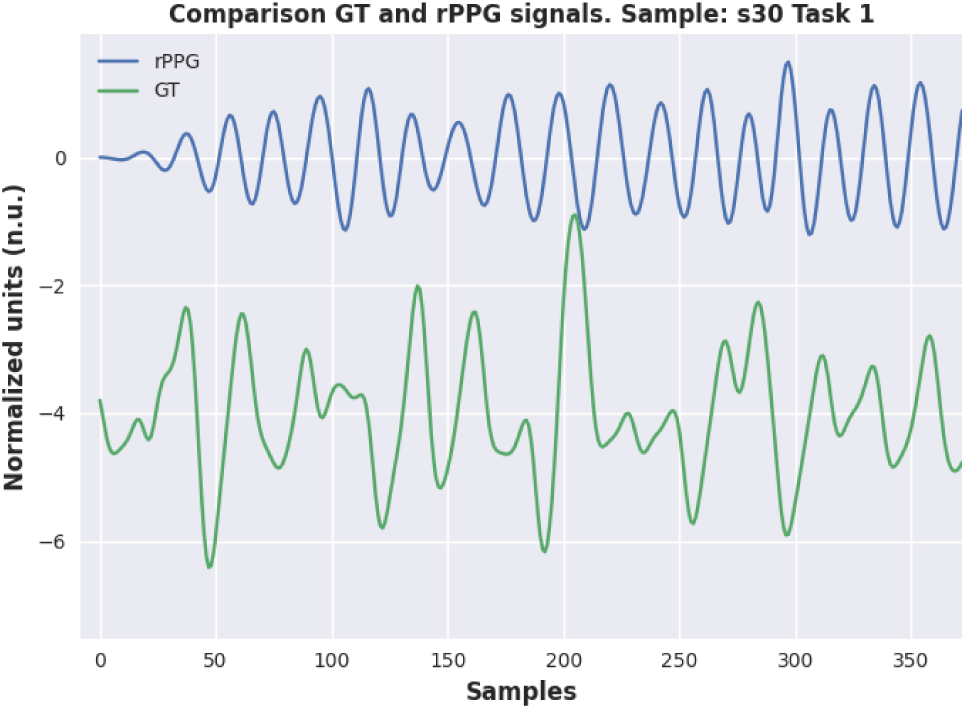
Example of the damaged signal.

Using the exclusion criteria based on the percentage of valid GT peaks > 60%, SNR > 4 dB, and a manual check, features from time, frequency, and non-linear domains were extracted from each one of the remaining samples using Contact and Remote signals after the post-processing algorithm. It also involves adaptive modeling techniques with the help of the Farr Institute (https://www.farrinstitute.org/healthcare-informatics).

From those HRV features, the most important ones, according to this work, are shown in Table IV, which demonstrates the average of each feature per task as well as the mean absolute error in the specific feature over all the remaining samples.

**Table IV:** Comparison of contact/remote features per task. Using proposed exclusion criteria.

| **T1** | Contact | Remote | MAE |
| --- | --- | --- | --- |
| HR (bpm) | 83.56 | 84.13 | 1.10 |
| IBI (ms) | 745.18 | 728.51 | 22.09 |
| RMSSD (ms) | 153.98 | 75.22 | 93.98 |
| SDNN (ms) | 127.53 | 77.40 | 62.10 |
| LF (s2) | 7.42 | 3.89 | 6.31 |
| HF (s2) | 6.29 | 1.85 | 5.18 |
| LF/HF | 1.40 | 2.13 | 0.98 |
| **T2** | Contact | Remote | MAE |
| HR (bpm) | 80.51 | 80.53 | 3.08 |
| IBI (ms) | 810.57 | 778.81 | 57.82 |
| RMSSD (ms) | 352.00 | 266.76 | 120.37 |
| SDNN (ms) | 267.14 | 212.93 | 88.64 |
| LF (s2) | 22.61 | 29.88 | 28.14 |
| HF (s2) | 23.77 | 14.39 | 13.07 |
| LF/HF | 0.97 | 1.40 | 0.73 |
| **T3** | Contact | Remote | MAE |
| HR (bpm) | 82.70 | 82.26 | 2.77 |
| IBI (ms) | 777.15 | 756.81 | 58.34 |
| RMSSD (ms) | 296.97 | 208.75 | 125.45 |
| SDNN (ms) | 224.09 | 165.73 | 89.28 |
| LF (s2) | 15.58 | 9.25 | 10.04 |
| HF (s2) | 17.46 | 9.69 | 11.76 |
| LF/HF | 1.09 | 1.05 | 0.52 |

Since the main purpose of the UBFC dataset was to create a database for social stress studies, in this work, we have investigated the ability of the proposed rPPG method to recognize stress states. First, statistical methods were used to explore the assumption that HRV tissues can differentiate between stress and non-stress states.

To investigate whether this assumption may be valid, we can visualize the distribution of certain HRV features across each task. Fig.13 shows the box plot distribution for each HRV feature across all 3 tasks.

**Figure 13:**
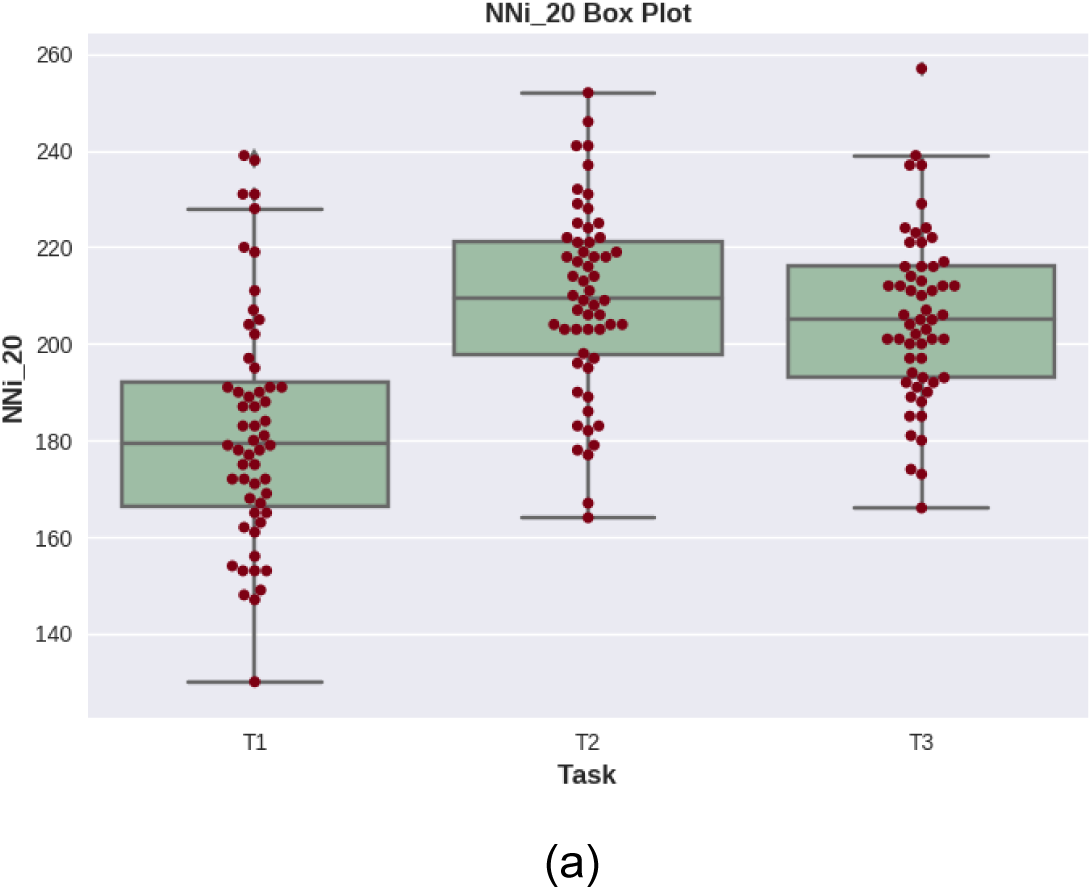
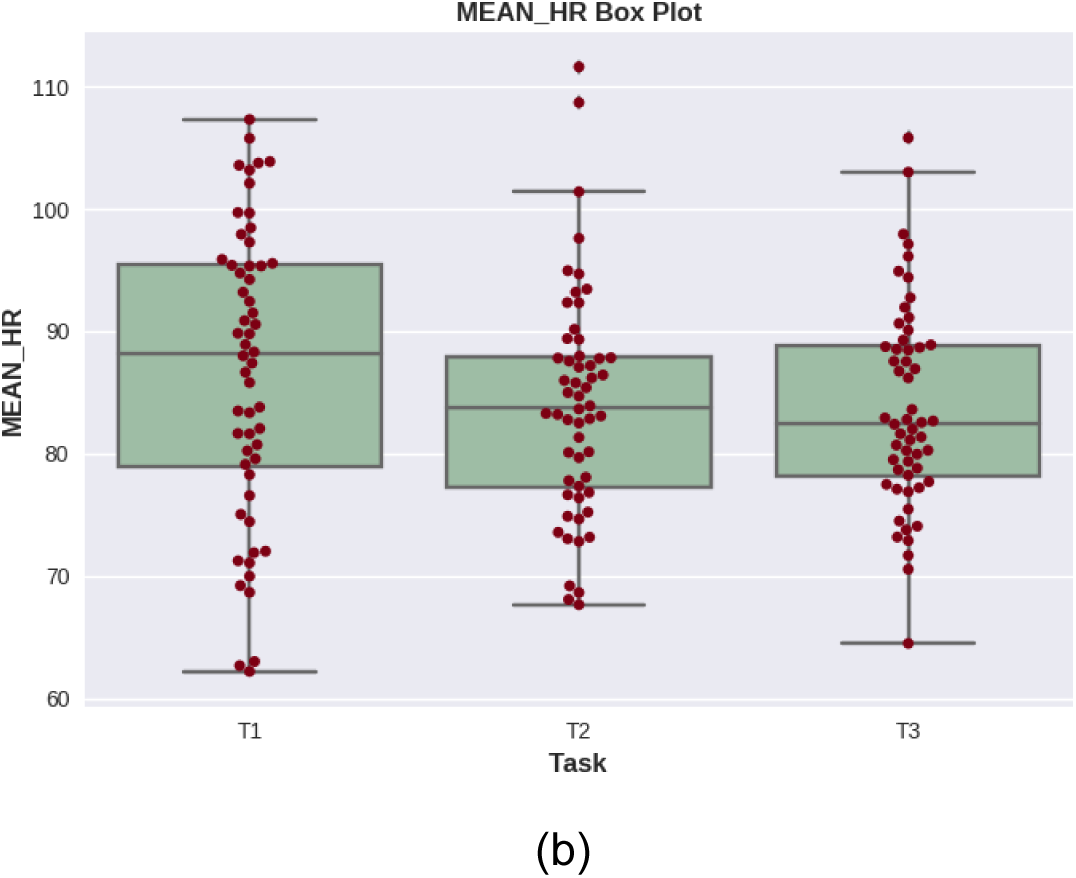
Box plot evaluation in each task for HRV features - a) NNi 20 and b) MEAN HR.

In addition, we investigated the use of machine learning models to correctly identify stress states and distinguish between tasks. Four classifiers were considered: Random Forest (RF) with 20 trees, Support Vector Machine (SVM) with a linear kernel, K-Nearest Neighbors (KNN) with k = 5, and Light Gradient Boosting Machine (Light-GBM).

Using metrics such as Accuracy, Precision, Sensitivity, Specificity, and F1-Score Table V shows the results for Stress State Recognition and Table VI shows the results for Task classification for each one of the models. In addition, Fig.14 shows the confusion matrix using the Random Forest model for each experiment.

**Table V:** Statistical analysis of this work, Stress Recognition.

| Model | Accuracy | Precision | Sensitivity | Specificity | F1-Score |
| --- | --- | --- | --- | --- | --- |
| **RF** | 83.11% | 77.08% | 94.87% | 71.05% | 85.05% |
| **SVM** | 72.72% | 68.00% | 87.17% | 57.89% | 76.40% |
| **KNN** | 71.42% | 68.08% | 82.05% | 60.52% | 74.41% |
| **LightGBM** | 77.92% | 70.37% | 97.43% | 57.89% | 81.72% |

**Table VI:** Statistical analysis of this work Task Recognition.

| Model | Accuracy | Precision | Sensitivity | Specificity | F1-Score |
| --- | --- | --- | --- | --- | --- |
| **RF** | 77.38% | 66.91% | 66.13% | 83.21% | 66.51% |
| **SVM** | 71.42% | 57.11% | 56.62% | 78.59% | 56.83% |
| **KNN** | 76.19% | 63.36% | 64.05% | 82.13% | 63.49% |
| **LightGBM** | 75.00% | 63.50% | 62.20% | 81.41% | 62.65% |

**Figure 14:**
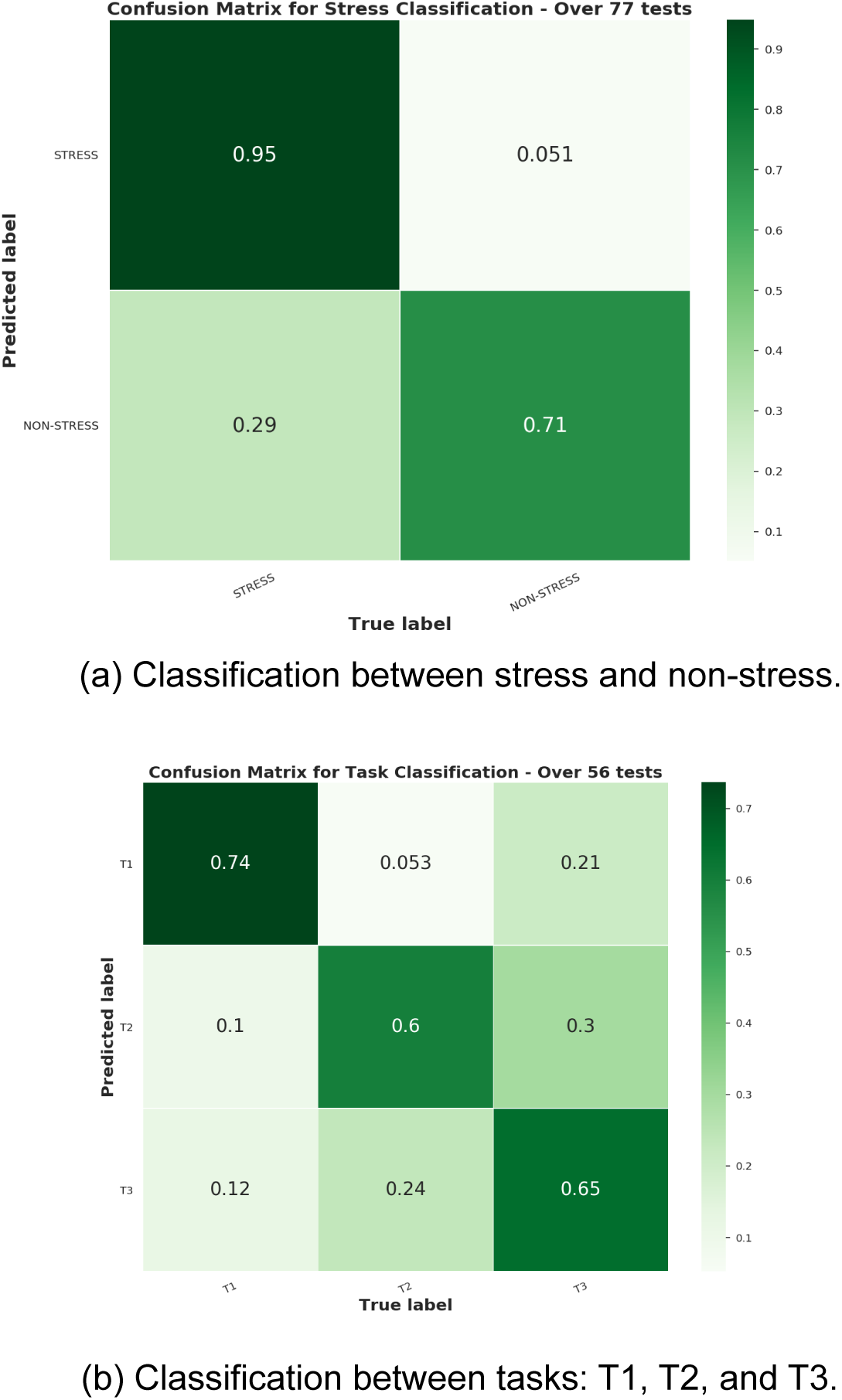
Confusion Matrix for Random Forest models.

## Discussion

In this work, we discovered that to accurately & fairly benchmark PPG signals against remote methods, there is a need to develop an exclusion criterion to evaluate which signals should be discarded and which signals should be included for comparative purposes. Across the UBFC dataset, we found that, on average, the quality of our rPPG signals was better than that of PPG signals from a contact sensor when applying the same evaluation metric to both.

After synchronizing the PPG and rPPG signals in time, we could analyze how well our rPPG method was able to measure ground truth pulsatile peaks in the signals and we use the F1 score as the metric for this evaluation. From Fig.6, it is clear to see that the rPPG and PPG signals were well synchronized in time, with a mean F1 score of above 90%, however, the lower F1 score for tasks T2 & T3 can be explained by some phase shift in the signals when motion is introduced.

Even though our post-processing algorithm can handle some damaged signals, as shown previously, in some samples, it is not possible to correct them to an acceptable level therefore the most appropriate approach is to filter out those files.

Table I details different thresholds that we can use for our exclusion criteria, which is based on the number of valid peaks in a signal as a percentage of all peaks detected in that signal. A threshold of 60% would mean that the signal must have at least 60% of its peaks marked as valid for comparative purposes. Table I shows how many files survived for each threshold and the comparative mean HR MAE as well as the minimum and maximum HR MAE.

However, despite applying this filter to the GT signals, we could still observe some outliers in terms of MAE values > 10 bpm, therefore to ensure that we are making a fair comparison on these samples, there was a need to include second exclusion criteria. Table II shows that while using a threshold of 60% in our first criteria and applying an additional SNR filter to the GT signal, there was a significant improvement in MAE values in both tasks T2 and T3.

There were still some outliers present in T1, therefore, to avoid comparing our rPPG signals against unacceptable GT signals, we performed a manual check on some of the GT samples. Eight GT signal samples from T1 were discarded due to their poor quality and none were further discarded from T2 & T3.

Overall, after applying various exclusion criteria to filter out poor quality GT signals, the HR MAE for T1, T2, and T3 was 1.10 bpm, 3.08 bpm, and 2.77 bpm respectively.

### Stress Recognition

To evaluate our ability to detect stress states, this work proposed the use of two different approaches, a statistically cal ANOVA test and machine learning.

Firstly, a one-way ANOVA was performed between subjects to compare the effect of each HRV characteristic sequentially on stress recognition. A probability p-value and a score F are defined to decide the significance of the variability. The p-value was compared to a significant level of 0.05. Hence, for p values lower than the significance level, it is possible to conclude that the group means are different, and the groups can be separable. However, before running the ANOVA test, we first need to check that the features are normally distributed using the Shapiro-Wilk test.

Based on the RR intervals from each signal, 32 HRV features were extracted. For those, there are 168 rows, which represents 56 subjects of 3 tasks. However, only four of those features are normally distributed according to the Shapiro-Wilk test: IBI, NNi 20, MEAN HR, and SAMPEN. The following conclusions can be made for those features.

The IBI and MEAN HR feature were not shown to be statistically significant for stress state at the p value < 0.05 level of p-value <0.05 for two conditions. IBI: [F(1, 166) = 1.75, p = 0.187], MEAN HR: [F(1, 166) = 1.00, p = 0.317]. However both NNi 20 and SAMPEN were shown to be statistically significant for stress state recognition at the p value < 0.05. NNi 20: [F(1, 166) = 8.65, p = 0.003], SAMPEN: [F(1, 166) = 7.34, p = 0.007]. We can conclude that both NNi 20 and SAMPEN were able to detect stress states with a 95% of confidence level.

These results suggest that in the time domain only the number of pairs of successive RR intervals that differ by more than 20 ms can differentiate between stress and non-stress. However, it should be noted that other HRV features could potentially accomplish this result.

In addition, the same experiment was conducted to find features that could differentiate between tasks.

Once again there was not a significant effect of MEAN HR on Task recognition at the p-value < 0.05 level for three conditions. MEAN HR: [F(2, 165) = 1.52, p = 0.221]. MEAN HR failed to separate tasks T1, T2, and T3.

However, there was a significant effect of IBI, NNi 20 and SAMPEN on Task recognition at the p value < 0.05 level for three conditions IBI: [F(2, 165) = 6.47, p = 0.001], NNi 20: [F(2, 165) = 26.97, p = 7.286e-11], SAM-PEN: [F(2, 165) = 8.27, p = 0.0003]. In fact, all of which, IBI, NNi 20 and SAMPEN succeeded a 95% confidence interval in separating at least two of three tasks T1, T2, and T3.

Even though ANOVA can statistically show whether the framework is able or not to separate the target, machine learning approaches are an easier way to classify whether the subject is under stress or not. The HRV features previously mentioned were extracted without any kind of windowing function, which means that there are only 168 entries that would be divided into train and test sets.

Besides that, the number of Stress and Non-Stress samples are heavily imbalanced, which might represent un- reliable results. To deal with this problem, the SMOTE algorithm proposed by [19] was applied to provide an evenly distributed dataset. This algorithm creates synthetic samples based on the Nearest Neighbors of the minority class sample. Hence, the number of entries increased to 232, equally distributed between Stress and Non-Stress classes.

Furthermore, as stated previously, the dataset has 32 HRV features; however, only 20 of them were used to build the models. A grid search analysis between globally assigned features was performed to define the best set of features.

Table V shows that all four ML models performed reasonably well. However, SVM, KNN, and LightGBM had problems with False Positives, represented by the Specificity metric. The Specificity around 60% can be explained by the situations where the subject was not under stress, however, the model classifies it as stress. In real-world situations, where a stress recognition system is applied to decide if a driver should rest, wrong classifications in the non-Stress class represent a more tolerable situation than wrong predictions in the Stress class. If the subject was under stress and the system allows him to drive, this wrong decision may contribute to a severe consequence such as a car accident.

Overall, the Random Forest model performed the best in most of the metrics and was able to achieve 83% ac- curacy with an F1-Score of 85%.

Moreover, Fig.14a shows that the RF model was able to achieve 95% of accuracy in the Stress class, however only 71% in classifying Non-Stress state.

Furthermore, in task classification the results were similar between the classifiers, however, the overall results were worse than in stress classification. The best result shows 77% accuracy and 66.51% F1-Score in the Random Forest model.

These results show that the model struggled to classify between T2 and T3, corroborating the hypothesis that T2 and T3 are similar tasks that should be within a single group, as activity.

### Conclusions

In this work, we proposed a benchmark comparison using the novel Multimodal UBFC dataset. The data set was built to allow studies on social stress since each of the 56 subjects was assigned to three different tasks, the first (T1) being a resting task and the second and third (T2 and T3) activity tasks. However, the main contribution of this dataset was to allow the comparison between Contact and Remote extraction of physiological information through BVP and rPPG signals.

The BVP signal had fewer filtering stages, which typically refers to a band-pass filter to remove unwanted frequencies as well as a moving average filter to smooth the signal. Whilst the rPPG signal was obtained using the method proposed by this work. Firstly, the RGB components were obtained from three different ROIs and averaged. Hence, a POS algorithm was used to mix the RGB channels into the rPPG signal, which ultimately was passed through a convolutional filter to improve the quality of the signal and make the peak detection more robust.

Furthermore, this work proposed a post-processing stage that seeks to filter out unreliable peaks. This method excludes peaks that do not abide by the IBI threshold rules. Although the post processing stage was possible to extract the percentage of validated peaks in each one of the signals as well as synchronize the contact and remote signals to measure the F1 score to evaluate how accurately our proposed method can detect pulsatile peaks.

The results showed that, according to the postprocessing algorithm, the rPPG signal had fewer corrupted peaks than the BVP signal, which represents a cleaner signal. Furthermore, by comparing contact and remote heart rate extracted using a 30-second sliding window approach there was a high correlation between the majority of the people. However, there were highly compromised samples, which correspond to signals with a very high level of noise that even the post-processing algorithm was unable to deal with.

This work performed an exclusion criterion to create a fair comparison using only reliable GT signals. Ultimately the exclusion criteria were based on the percentage of validated GT peaks above 60%, SNR of GT signal above 4 dB, and a manual GT signal quality check. The results have shown a considerable improvement, with 1.10 bpm of mean error in T1 in 47 samples, bpm in T2 in 22 samples, and finally 2.77 bpm in T3 in 34 samples.

In terms of stress recognition, this work proposed two different approaches. The ANOVA test was conducted over HRV features extracted from the rPPG signal, results show that two features were able to statistically differentiate between stress and non-stress. Moreover, three HRV features were able to differentiate between tasks. Using Tukey’s test, we discovered that those three characteristics were able to differentiate between T1 (rest task) and T2 or T3, which represents the tasks for which there was a higher level of activity.

Ultimately, four machine learning models were evaluated to investigate if any of them were able to classify between stress and non-stress as well as between T1, T2, and T3. Results have shown that the Random Forest model reached 83.11% of accuracy, 77.08% of precision, and 94.87% of recall in the binary classification, which denotes the effectiveness of the model in classifying the Stress status.

In task recognition problems, once again the Random Forest model obtained the best score with 77.38% of ac- curacy, 66.91% of precision, and 66.13% of recall. Although these results highlighted the difficulty of distinguishing between T2 and T3, it also showed that the model was able to differentiate between T1 and T2 or T3.

Further work will include the enhancement of the rPPG core algorithm to better deal with motion robustness, once the method can improve the results in those noisy signals it will considerably decrease the mean error. Moreover, further studies in stress recognition using the remote signal will be performed, through the results of this work can be concluded that there is a correlation between HRV features and stress tasks, however, with additional data, it will be possible to conclude which features are more likely to carry the stress information.

## Data Availability

Researchers interested in access to the data may contact Nikhil Sehgal at, also see https://vastmindz.com/contact/. The authors will assist with any negotiable data use agreements and gain access to the data for any replication efforts following publication.

## Acknowledgment

We express our gratitude to Professor Mel Lobo for his contributions from an academic/clinical perspective. We also express our thanks to Farr Institute (https://www.farrinstitute.org/healthcare-informatics/) for their support in healthcare informatics.

## Abbreviations

AI: Artificial Intelligence
ANOVA: Analysis of Variance
ANN: Artificial neural network
ANS: Autonomic Nervous System
BVP: Blood Volume Pulse
CVD: Cardiovascular diseases
HRV: Heart Rate Variability
POS: Plane-Orthogonal-to-Skin
PPG: photoplethysmography
rPPG: Remote Photoplethysmography
SNR: signal-to-noise ratio
WHO: World Health Organization

## References

1. World Health Organisation. 2021. Cardiovascular diseases (CVDs). [online] Available at: https://www.who.int/news-room/fact-sheets/detail/cardiovascular-diseases-(CVDS). Accessed 20 June 2021.

2. American Institute of Stress, “42 worrying workplace stress statistics,” 2021, accessed on 20 June 2021. [Online]. Available: https://www.stress.org/42-worrying-workplace-stress-statistics

3. H. Kim, E. Cheon, D. Bai, Y. Lee, and B. Koo, “Stress and heart rate variability: A meta-analysis and review of the literature,” Korean Neuropsychiatric Association, 2018.

4. P. Viola and M. Jones, “robust real-time object detection”, International Journal of Computer Vision, 2001.

5. D. McDuff, S. Gontarek, and R. W. Picard, “Improvements in remote cardiopulmonary measurement using a five-band digital camera,” IEEE Transactions on Biomedical Engineering, vol. 61, no. 10, pp. 2593–2601, 2014.

6. M. Lewandowska, J. Rumin’ski, T. Kocejko, and J. Nowak, “Measuring pulse rate with a webcam - a non-contact method for evaluating cardiac activity,” in 2011 Federated Conference on Computer Science and Information Systems (FedCSIS), 2011, pp. 405–410.

7. G. de Haan and V. Jeanne, “robust pulse rate from chrominance-based rppg”,” IEEE Transactions on Biomedical Engineering, vol. 60, no. 10, pp. 2878–2886, 2013.

8. W. Wang, A. C. den Brinker, S. Stuijk, and G. de Haan, “Algorithmic principles of remote ppg,” IEEE Transactions on Biomedical Engineering, vol. 64, no. 7, pp. 1479–1491, 2017.

9. R. Meziati, Y. Benezeth, P. Oliveira, J. Chappé, and F. Yang, “Ubfc-phys,” 2021. [Online]. Available: https://dx.doi.org/10.21227/5da0-7344

10. A multimodal database for psychophysiological studies of social stress”,” IEEE Transactions on Affective Computing, 2021.

11. A. Savitzky and M. J. E. Golay, “Smoothing and differentiation of data by simplified least squares procedures.” American Chemical Society, 1964.

12. I. Selesnick and C. Burrus, “Generalized digital butter-worth filter design,” IEEE Transactions on Signal Processing, vol. 46, no. 6, pp. 1688–1694, 1998.

13. G. Bradski, “The OpenCV Library,” Dr. Dobb’s Journal of Software Tools, 2000.

14. E. Ostertagova and O. Ostertag, “Methodology and application of one-way ANOVA,” American Journal of Mechanical Engineering, vol. 1, pp. 256–261, 11 2013.

15. D. C. Montgomery and G. C. Runger, Applied statistics and probability for engineers. John Wiley and Sons, 2013.

16. S. S. Shapiro and M. B. Wilk, “An analysis of variance test for normality (complete samples),” Biometrika, vol. 52, no. 3-4, pp. 591–611, Dec 1965. [Online]. Available: https://doi.org/10.1093/biomet/52.3-4.591

17. B. Barton and J. Peat, Medical statistics: A guide to SPSS, data analysis and critical appraisal, 2nd ed. John Wiley and Sons, 2014.

18. J. W. Tukey, Exploratory Data Analysis. Pearson, 1977.

19. N. V. Chawla, K. W. Bowyer, L. O. Hall, and W. P. Kegelmeyer, “Smote: Synthetic minority over-sampling technique,” J. Artif. Int. Res., vol. 16, no. 1, p. 321–357, Jun. 2002.

